# Stress and Sleep Duration in Immune and Neuroendocrine Patterning. An Analytical Triangulation in ELSA

**DOI:** 10.1101/2024.07.23.24310898

**Authors:** Odessa S. Hamilton, Andrew Steptoe

**Affiliations:** Department of Behavioural Science and Health, Institute of Epidemiology and Health Care, University College London, 1-19 Torrington Place, London WC1E 7HB, UK

**Author notes:** **Corresponding Author:** Odessa S. Hamilton, Department of Behavioural Science and Health, Institute of Epidemiology and Health Care, University College London, 1-19 Torrington Place, London WC1E 7HB, UK.

**Keywords:** Financial Stress, Sleep Duration, Inflammation, Longitudinal, Genetic Risk, Latent Profile Analysis

## Abstract

**Background**. Proinflammatory and neuroendocrine mediators are implicated in disease aetiopathogenesis. Stress increases concentrations of immune-neuroendocrine biomarkers through a complex network of brain-body signalling pathways. Suboptimal sleep further modulates these processes by altering major effector systems that sensitise the brain to stress. Given the ubiquitous, impactful nature of material deprivation, we tested for a synergistic association of financial stress and suboptimal sleep with these molecular processes. **Methods**. With data drawn from the English Longitudinal Study of Ageing (ELSA), associations were tested on 4,940 participants (∼66±9.4 years) across four-years (2008-2012). Through analytical triangulation, we tested whether financial stress (>60% insufficient resources) and suboptimal sleep (≤5/≥9 hours) were independently and interactively associated with immune-neuroendocrine profiles, derived from a latent profile analysis (LPA) of C-reactive protein, fibrinogen, white blood cell counts, hair cortisol, and insulin-like growth factor-1. **Results**. A three-class LPA model offered the greatest parsimony. After adjustment for genetic predisposition, sociodemographics, lifestyle, and health, financial stress was associated with short-sleep cross-sectionally (RRR=1.45; 95%CI=1.18-1.79; *p*<0.001) and longitudinally (RRR=1.31; 95%CI=1.02-1.68; *p*=0.035), and it increased risk of belonging to the *high-risk* biomarker profile by 42% (95%CI=1.12-1.80; *p*=0.004). Suboptimal sleep was not related to future risk of *high-risk* profile membership, nor did it moderate financial stress-biomarker profile associations. **Discussion**. Results advance psychoneuroimmunological knowledge by revealing how immune-neuroendocrine markers cluster in older cohorts and respond to financial stress over time. Financial stress associations with short-sleep are supported. The null role of suboptimal sleep, as exposure and mediator, in profile membership, provides valuable insight into the dynamic role of sleep in immune-neuroendocrine processes.

## Introduction

The most intriguing aspect of inflammation is the plurality of its modulators and its breadth of effects that support health or contribute to morbidity, both physical and mental, and mortality worldwide (Jerison, 2024; Furman et al., 2019). With multilateral, cross-blood-brain-barrier signalling between the immune, nervous, and endocrine systems (Becher et al., 2017), avenues of inquiry have found that abnormal circulating concentrations of pro-inflammatory and endocrine-mediated biomarkers have been implicated in the genesis and progression of disease and psychiatric disorder across a spectrum of severity (Furman et al., 2019; Hamilton & Steptoe, 2022; Tomasik et al., 2023; Yuan et al., 2019). A disease burden that not only intimates premature death but suffering as people live with it (Whiteford et al., 2013).

Human, animal, and *in vitro* studies converge to the understanding that exogenous acute and chronic stress directs adverse effects on immunologic mechanisms (Bierhaus et al., 2006), with material deprivation ubiquitous and among the strongest indicators of stress (Hamilton, Iob, et al., 2023; McEwen, 2007; Muscatell et al., 2020). A stress modulated hypothalamic–pituitary-adrenal axis (HPA-axis) orchestrates a complex network of brain-body responses that increase circulating concentrations of immune and neuroendocrine mediators, with serious consequences to health (Bierhaus et al., 2006; Steptoe et al., 2007). Various mechanisms may underlie this response, from stress-induced reductions in plasma volume, upregulation of synthesis, or expansion of the cell pool contributing to synthesis (Steptoe et al., 2007).

When sleep deprived, alterations can also be seen to major effector systems, including the HPA-axis and the sympathetic nervous system (SNS). This corresponds to catecholamines elevations that drive abnormal inflammatory responses, with shifts seen to the levels and temporal profile of the response, and functional alterations in the expression of pro-inflammatory cytokines that then interact with the brain through humoral, neural, and cellular pathways (Garbarino et al., 2021; Irwin, 2006, 2019). Short sleep trajectories are particularly relevant to inflammatory responses (Bakour et al., 2017); a more salient concern to cohorts prone to age-related declines in sleep efficiency (Kocevska et al., 2020). Even one night of sleep restricted to 4-hours can led to over a three-fold increase monocyte production of interleukin-6 (IL-6), tumor necrosis factor-alpha (TNF-α), and messenger RNA (mRNAs; (Irwin, 2006). While a single hour of shorter sleep has been associated with CRP and IL-6 elevations (Ferrie et al., 2013). In an RCT, sleep-deprived adults had higher baseline cortisol levels and an exaggerated cortisol response to stress than well-rested adults (Minkel et al., 2014). Findings that suggest that sleep deprivation contribute to inflammatory processes by sensitising the brain to stress (Uy et al., 2022). But a meta-analytic review (*n*>50,000 participants) found that neither total nor partial sleep deprivation reliably increased inflammation, with results greatly depending on the sleep index (Irwin et al., 2016).

It is equally important to consider whether stress is as antithetical to good sleep as poor sleep is to stress, with evidence that they co-occur and are reciprocally reinforcing (Jones & Gatchel, 2018). Stress has been implicated as a predisposing factor for shorter actigraphy-determined total sleep time, with worse sleep associated with greater stress perception (Slavish et al., 2021). In another study, a one unit increase in self-reported stress was associated with a 3min decrease in actigraphic and self-reported total sleep, with no significant associations between sleep parameters and next-day stress (Yap et al., 2020). Results have been difficult to unravel, with inconsistencies between and within studies. Findings have largely depended on the type, acuteness and chronicity of stress, or the degree of sleep abnormality and whether it is assessed by self-report, actigraphy, or polysomnography (Beck et al., 2022; Slavish et al., 2021).

A better understanding of the interaction between financial-related stress and suboptimal sleep may elucidate how they independently and collectively influence immune-neuroendocrine biomarkers as a clinically relevant pathway to disease. It is possible that the impact of experiencing both is synergistic rather than additive or multiplicative, reflecting worse combined effects on biological processes. This raises the question of whether the nature of the relationship between financial stress and inflammatory processes is altered by the presence of suboptimal sleep, having controlled for genetic predisposition and a rich selection of confounders.

When results from different approaches, across disciplines, with unrelated source of bias, point to the same conclusion it strengthens confidence in the finding (Lawlor et al., 2017). For this reason, in this study, we leverage several statistical, genomic, and epidemiological techniques to address this research question. First, a latent profile analysis (LPA) was used to capture the heterogeneity and latent structure of five immune-neuroendocrine biomarkers (i.e., C-reactive protein [CRP]; fibrinogen [Fb]; white blood cell counts [WBCC]; hair cortisol [Cortisol]; and insulin-like growth-factor-1 [IGF-1]), which decomposed the population into a small number of groups (Bauer, 2005). Second, polygenic scores (PGS) were taken into account in the analyses to index genetic predisposition (Hamilton, Steptoe, et al., 2023) and to understand genetic risk for immune-neuroendocrine profile membership. Third, the construction of a directed acyclic graph (DAG) served to identify sources of bias and visually represent the complex causal pathways between variables (Lipsky & Greenland, 2022). Fourth, an observational, longitudinal design allowed for temporal changes to be traced in a way that points to directionality. Finally, effect modification between financial stress and suboptimal sleep was evaluated to uncover subgroup differences in patterning of immune-neuroendocrine processes. We hypothesised that financial stress and suboptimal sleep would be independently associated with belonging to the highest risk latent profile of immune-neuroendocrine biomarkers. Separately, we anticipated the presence of an interaction between financial stress and suboptimal sleep in this association.

## METHODS

### Study Design

Fully anonymised data were drawn from the English Longitudinal Study of Ageing (ELSA), an ongoing multidisciplinary, observational study, with a longitudinal response rate of 70% (Steptoe et al., 2013). In line with the National Census, ELSA is representative of the non-institutionalised general population aged ≥50 in England. Data collection is performed in participants’ homes, via computer-assisted personal interviews (CAPI) and self-completed questionnaires biennially, then nurse visits quadrennially to collect biological samples. All participants provide written consent and ethical approval was granted by the National Research Ethics Service. Here, longitudinal sample derivation is from wave 4 (2008) and wave 6 (2012). Analyses were weighted using longitudinal survey weights. A total of 6,523 participants had complete measures and at least one biomarker at baseline. After exclusions of CRP values >20 mg/L (*n*=116), the sample was 6,407. Of these, 1,467 had missing genetic data, leaving an analytic sample of 4,940 (Figure Supplementary [S] 1).

### Exposure

Financial stress was indexed by financial strain, a binary measure of the perceived chance of not having enough financial resources in the future to meet needs (categorised by 0; 1-39; 40-60; 61-99; 100% and dichotomised at >60%). The higher the percentage, the higher the belief of having insufficient resources and, thus, the higher the stress experience.

### Moderator

Sleep duration was measured with an open-ended question, asking participants about the length of their sleep on an average weeknight. Outlier values greater than three standard deviations (±) from the mean (M) were excluded from the raw data. To improve model fit, interpretability, and to avoid non-linear interaction complexity, sleep duration was demarcated by “≤5 hrs” (i.e., short-sleep), “>5-<9 hrs” (i.e., optimal-sleep), and “≥9 hrs” (i.e., long-sleep), categories that align with earlier research (Hamilton, Steptoe, et al., 2023).

### Outcomes

Immune-neuroendocrine biomarkers entered into the Latent Profile Analysis (LPA) included high-sensitivity plasma C-reactive protein (CRP; mg/L), plasma fibrinogen (Fb; g/L), white blood cell counts (WBCC; 10^9^/L), hair cortisol (cortisol; pg/mg), and serum insulin-like growth factor-1 (IGF-1; mmol/L). Exclusion criteria for bloods included coagulation and haematological disorders, being on anticoagulant medication or having a history of convulsions. Blood samples were discarded if deemed insufficient or unsuitable (e.g., haemolysed; received >5 days post-collection). Collection details can be found in Supplementary Materials (SM) 1.

### Covariates

Covariates were built into the model *a priori,* on the basis of a DAG (SM2). The DAG was developed to trace causal pathways, mitigate bias, optimise parameter inferences, and improve estimate accuracy. This epidemiological approach led variables, likely on the causal pathway between stress, suboptimal sleep, and immune-neuroendocrine processes, to be excluded from the main models, because conditioning on them would have introduced collider bias (Hernán & Monge, 2023). All measured at baseline, covariates included *demographics*: age (≥50 years); age^2^ ([squared] to account for non-linearity); sex (male/female); *genetic variables*: 10 PCs, PGS for CRP, WBCC, Cortisol, IGF-1, and sleep duration; *socioeconomic variables*: education (higher; lower; or none); wealth (total net household wealth, as determined by the summation of property [minus mortgage], business assets, possessions, liquid assets; cash, savings, investments, artwork, and jewellery, net of debt, exclusive of pension wealth); *lifestyle variables*: smoking status (non-smokers and ex-smokers/smokers); weekly alcohol consumption (low <3/high ≥3 day); weekly physical activity (sedentary and moderate/vigorous activity); *health variables*: limiting longstanding illness (an enduring condition that limits activity); any self-reported clinician diagnosis of abnormal heart rhythm; angina; arthritis; asthma; cancer; chronic lung disease; congestive heart failure; coronary heart disease; diabetes; heart murmur; hypertension; osteoporosis; dementia; Parkinson’s disease; or psychiatric disorder; difficulty with mobility (walking 100 yards; sitting 2-hours; rising from chairs after sitting long periods; climbing stairs; stooping, kneeling, or crouching; reaching or extending arms above shoulders; pulling or pushing large objects; lifting or carrying objects over 10lb; picking-up a 5p coin).

### Statistical Analyses

#### Polygenic Scores (PGS)

The derivation of the PGSs for CRP, WBCC, IGF-1, Cortisol, and sleep duration has been detailed in the supplementary (SM3). Z-scores were used to standardise all PGSs, so values from different distributions could be equitably compared and so interpretation could be improved. The genome-wide genotyping, funded by the Economic and Social Research Council (ESRC), was performed at University College London (UCL) Genomics in 2013-2014 using the llumina HumanOmni2.5 BeadChips (HumanOmni2.5-4v1, HumanOmni2.5-8v1.3).

#### Multiple Imputation

Owing to attrition and item non-response missingness ranged 0.00-61.0% (Table S1; SM4). Imputation was performed using missForest (RStudio v.2022.02.2); a Random Forest algorithm-based machine learning imputation method that builds decision trees during training and combines predictions iteratively until convergence to ensure accurate predictions with minimal overfitting (Stekhoven & Bühlmann, 2012). Imputation yielded a minimal error for continuous variables (Normalized Root Mean Squared Error [NRMSE]=0.07%) and categorical variables (Proportion of Falsely Classified [PFC]=0.05%).

#### Latent Profile Analysis (LPA)

The LPA was conducted to cluster the sample according to concentrations of immune-neuroendocrine biomarkers; CRP, Fb, WBCC, Cortisol, and IGF-1. An LPA model for observed variable *A* that can be expressed as:

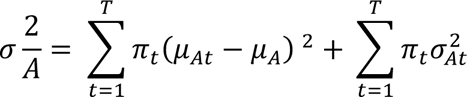

where *µ_At_* and 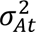 denote (*t*) class-specific means and variances for variable *A*, and *π_t_* show the proportion of *N* participants that belong to class *t*. A stepwise approach was taken to identify the optimal number of latent profiles and the number of profiles was determined on the basis of multiple parameters that have been detailed in the supplementary (SM5). Once the number of latent profiles was established, individuals within the sample were assigned to a cluster for which they had the greatest posterior probability of belonging to.

#### Association Analyses

Baseline characteristics were expressed as means and proportions, with Analysis of Variance (ANOVA) and Chi-squared (χ^2^) comparisons on biomarkers. Logarithmic transformation was performed on CRP, WBCC, Cortisol, and IGF-1 values because of their originally skewed distribution, but Fb was normally distributed. There was no evidence of attrition bias due to systematic differences in missing data or differential loss-to-follow-up (Table S1; Figure S1). Multivariable, multinomial regressions were used to test several key associations. First, the cross-sectional association between financial stress and suboptimal sleep at wave (W) 4 (2008). Second, the longitudinal association of financial stress at W4 on suboptimal sleep, then immune-neuroendocrine profile membership at W6 (2012). Third, the association between suboptimal sleep at W4 and immune-neuroendocrine profile membership at W6. Finally, a multiplicative interaction term was applied between financial stress and suboptimal sleep at W4 on immune-neuroendocrine profiles at W6; expressed by an ordinary least squares (OLS) regression equation:

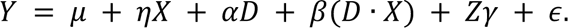

where ***Y*** represents immune-neuroendocrine profile membership (W6 *outcome*); ***D*** represents financial stress (W4 *exposure*); ***X*** represents suboptimal sleep (W4 *moderator*); ***D***·***X*** is the interaction term with its constituent first-order terms (***D*** and ***X***); ***Z*** is a vector of covariates, while *µ* and *̅* represent the constant and error terms, respectively. The magnitude of the interaction coefficient represents the estimated change in the effect of the focal exposure ***D*** on the outcome ***Y*** for a single unit change in the moderator ***X***. All regression assumptions were met (Aguinis & Gottfredson, 2010; Hainmueller et al., 2018). The highest risk category was reported against the lowest risk reference. Results were reported as relative risk ratios (RRR), with standard errors (SE) and 95% confidence intervals (95% CI). Analyses were two-tailed. Different models were fitted to understand the role of covariates on associations: Model (M) 1 was unadjusted; M2 adjusted for baseline immune-neuroendocrine profiles; M3 controlled for demographic and genetic variables; M4 was fully adjusted. Association analyses were conducted in Stata 18.1 (STATA CorpLP, USA).

#### Sensitivity Analyses

To test the robustness of the results, five additional analyses were performed. First, owing to known changes in stress perception (Graham et al., 2006) and sleep trajectories as we age (Kocevska et al., 2020), it is conceivable that these factors are a less salient risk to biological processes in later adulthood, so associations were stratified by median age (≥65). Second, sex-stratified analyses were performed because individuals who experience short sleep are more likely to be men, while women are more likely to experience long sleep (Bakour et al., 2017; Prather et al., 2015), with sex differences also reported in stress experience (Bale & Epperson, 2015). Third, excess adipose tissue, as measured by BMI, is a likely mediator because of its shared genetic basis with suboptimal sleep (Garfield et al., 2019), along with its known roles in stress (Wardle et al., 2011) and inflammation (Brydon et al., 2008), so independent analyses controlled for BMI. Fourth, caseness of suboptimal sleep durations were low, so the thresholds were changed to “≤6 hr” (i.e., short-sleep), “>6-<8 hr” (i.e., optimal-sleep), and “≥8 hr” (i.e., long-sleep) to ensure that results were not contingent on power nor the extremities of short and long sleep. Fifth, PGSs can detect a common genetic basis between traits and provide a prediction of an individual’s genetic risk for a particular disease, or in this case a biological outcome (Pain et al., 2022). PGSs for short and long sleep were, therefore, used to test associations with immune-neuroendocrine profile membership, for results less encumbered by confounding, and to provide stronger evidence of a possible causal effect between the two. There are no GWASs from which a PGS for stress could be derived.

## RESULTS

### Cohort characteristics

Sample characteristics are described in Table 1. The sample comprised 4,940 (Figure S1) from whom total baseline data were available. Participants were male (45.3%) and female (54.7%), aged 66.3 years on average (±9.35; range50-99). Of these, 17.0% experienced financial strain, 12.7% short sleep, and 1.7% long sleep. Although individual trajectories varied widely, biomarkers were relatively stable from baseline to follow-up. CRP was linearly correlated with Fb (*r*=0.707); WBCC (*r*=0.448); Cortisol (*r*=0.281); and IGF-1 (*r*=-0.167; all *p*=<0.001). All other correlations can be found in Table S2. Participants were, on average, overweight (73.4%), but active (72.9%), most consumed alcohol less than three days a week (64.3%) and were non-smokers (87.3%). The majority were without disorder (67.2%), and most had no limiting longstanding illness (68.5%), but similar proportions were with and without mobility difficulties (45.8/54.2%).

**Table 1.** Sample characteristics.

| Variable |  | Baseline (N= 4,940) |  |  |  |
| --- | --- | --- | --- | --- | --- |
| | | N / M (SD) | % / Range | ANOVA | $\chi^2$ |
| Age |  | 66.3 (9.35) | 50-99 | <0.001 |  |
| Age (Binary) | <65 Md | 2,436 | 49.31 |  | <0.001 |
|  | ≥65 Md | 2,504 | 50.69 |  |  |
| Sex | Male | 2,237 | 45.3 |  | 0.073 |
|  | Female | 2,703 | 54.7 |  |  |
| Education | Higher | 1,589 | 32.2 |  | <0.001 |
|  | Lower | 1,543 | 31.2 |  |  |
|  | Alternative/No | 1,808 | 36.6 |  |  |
| Wealth | Lowest | 1,573 | 31.8 |  | <0.001 |
|  | Middle | 2,014 | 40.8 |  |  |
|  | Highest | 1,353 | 27.4 |  |  |
| Smoking Status | Never/Ex-Smokers | 4,312 | 87.3 |  | <0.001 |
|  | Current Smoker | 628 | 12.7 |  |  |
| Alcohol Consumption | <3 days a week | 3,175 | 64.3 |  | <0.001 |
|  | ≥3 days a week | 1,765 | 35.7 |  |  |
| Physical Activity | Sedentary | 1,340 | 27.1 |  | <0.001 |
|  | Active | 3,600 | 72.9 |  |  |
| Mobility | Mobile | 2,678 | 54.2 |  | <0.001 |
|  | Not Mobile | 2,262 | 45.8 |  |  |
| Limiting Longstanding Illness | None | 3,386 | 68.5 |  | <0.001 |
|  | Present | 1,554 | 31.5 |  |  |
| Health | No health condition | 3,319 | 67.2 |  | <0.001 |
|  | At least one health condition | 1,621 | 32.8 |  |  |
| BMI | <25, Underweight/Normal | 1,312 | 26.6 |  | <0.001 |
|  | 25-30, Overweight: Pre-obese | 2,213 | 44.8 |  |  |
|  | 30 or over, Obese | 1,415 | 28.6 |  |  |
| PGS for CRP |  | 0.00 (1.00) | -3.50 (3.76) | 0.235 |  |
| PGS for WBCC |  | 0.00 (1.00) | -4.03 (2.58) | 0.123 |  |
| PGS for IGF-1 |  | 0.00 (1.00) | -3.69 (3.83) | 0.411 |  |
| PGS for Cortisol |  | 0.00 (1.00) | -3.47 (3.28) | 0.928 |  |
| PGS for Sleep Duration |  | 0.00 (1.00) | -4.09 (2.87) | 0.449 |  |
| CRP* (mg/L; Baseline) |  | 0.28 (0.46) | -0.70-1.30 | <0.001 |  |
| CRP* (mg/L; Follow-up) |  | 0.40 (0.48) | -1.00-1.30 | <0.001 |  |
| Fb (g/L; Baseline) |  | 3.38 (.56) | 1.30-5.90 | <0.001 |  |
| Fb (g/L; Follow-up) |  | 3.12 (0.52) | 1.50-5.80 | <0.001 |  |
| WBCC* (nmol/L; Baseline) |  | 0.79 (0.13) | -0.10-1.50 | 0.062 |  |
| WBCC* (nmol/L; Follow-up) |  | 0.81 (0.11) | 0.34-1.51 | <0.001 |  |
| IGF-1* (nmol/L; Baseline) |  | 1.18 (0.16) | 0.30-1.81 | <0.001 |  |
| IGF-1* (nmol/L; Follow-up) |  | 1.18 (0.13) | 0.60-1.76 | <0.001 |  |
| Cortisol* (nmol/L; Follow-up) |  | 1.23 (0.65) | -0.85-2.82 | 0.158 |  |
| Stress (indexed by Financial Strain) | No Strain (0-60%) | 4,099 | 83.0 |  | <0.001 |
|  | Strain (61-100%) | 841 | 17.0 |  |  |
| Sleep Duration | Short Sleep | 627 | 12.7 |  | <0.001 |
|  | Optimal Sleep | 4,227 | 85.6 |  |  |
|  | Long Sleep | 86 | 1.7 |  |  |
| I-N Profiles (Baseline) | *Low-risk* | 2,590 | 52.4 |  | <0.001 |
|  | *Moderate-risk* | 1,773 | 35.9 |  |  |
|  | *High-risk* | 577 | 11.7 |  |  |
| I-N Profiles (Follow-up) | *Low-risk* | 1,739 | 35.2 |  | - |
|  | *Moderate-risk* | 1,951 | 39.5 |  |  |
|  | *High-risk* | 1,250 | 25.3 |  |  |

Notes: ELSA, waves 4-6 (2008-2012); N = observations; M = Mean; Md = Median; % = percentage frequencies; SD = standard deviations; ANOVA = Analysis of Variance between the exposed and unexposed for continuous variables with a 3-level categorical variable; $\chi^2$ = Pearson Chi square test significance between the exposed and unexposed for categorical variables with a 3-level categorical variable; < = less than; ≥ = greater than or equal to; BMI = Body Mass Index; PGS = Polygenic Score; CRP = C-reactive protein; Fb = Fibrinogen; WBCC = White Blood Cell Counts; IGF-1 = Insulin-growth factor-1; Cortisol = Hair Cortisol; \* Log-transformed variable; I-N = Immune and Neuroendocrine.

### Latent profile analysis of immune and neuroendocrine biomarkers

A three-profile model of W6 immune-neuroendocrine biomarkers provided the most parsimonious fit to the data (Table S3; Figures S4 [a-g]). After which there were limited returns in AIC and BIC value (Figure S5), entropy was 0.67 (Figure S6), the mean posterior probabilities did not exceed 0.70, each profile had ≥5% of participants (Figure S7); and there was theoretical meaning to the profiles. Profile 1, defined as ‘*low-risk*’, included 35.2% of the sample (M_age_=64.05; ±7.72; _male_47.44/_female_52.56%), characterised by individuals with low CRP, Fb, WBCC, Cortisol, and high IGF-1. The modal profile 2, defined as ‘*moderate-risk*’, included 39.5% of the sample (M_age_=66.53; ±9.31; _male_43.77/_female_56.23%) and consisted of individuals with moderate CRP, Fb, WBCC, Cortisol, and IGF-1 levels. Finally, profile 3, defined as ‘*high-risk*’, included 25.3% of the sample (M_age_=69.13; ±10.60; _male_44.68/_female_55.32%;) and was marked by a high probability of high CRP, Fb, WBCC, Cortisol, and low IGF-1 (Figure 6). The number of individuals within the *low-risk* group fell by 17.2% from baseline (W4; 2008) to follow-up (W6; 2012), with the *moderate-risk* group increasing by 3.6%, and the *high-risk* group increasing by 13.6% (Table 1).

### Cross-sectional associations of financial stress with suboptimal sleep

As shown in Table 2, after adjustment for, age, sex, and genetic predisposition to adverse immune-neuroendocrine biological signatures, the experience of financial-related stress was cross-sectionally associated with an 80% greater risk of experiencing short sleep (RRR=1.80; 95%CI=1.47-2.20; *p*<0.001), but not long sleep (RRR=1.24; 95%CI=0.70-2.19; *p*=0.463). And associations held after full adjustment, with financial stress being associated with a 45% greater risk of short sleep (RRR=1.45; 95%CI=1.18-1.79; *p*<0.001).

**Table 2.** Cross-sectional associations between stress and suboptimal sleep.

| Adjustments | Stress |  |  |  |  |
| --- | --- | --- | --- | --- | --- |
|  | RRR | SE | 95% CI |  | *p* |
| Short Sleep |  |  |  |  |  |
| Model 1: *Unadjusted* | 1.80 | 0.18 | 1.47 | 2.19 | <0.001 |
| Model 2: *Model 2 + demographics & genetics* <sup>a, b</sup> | 1.80 | 0.19 | 1.47 | 2.20 | <0.001 |
| Model 3: *Fully Adjusted* <sup>c</sup> | 1.45 | 0.15 | 1.18 | 1.79 | <0.001 |
| Long Sleep |  |  |  |  |  |
| Model 1: *Unadjusted* | 1.13 | 0.32 | 0.64 | 1.98 | 0.680 |
| Model 2: *Model 2 + demographics & genetics* <sup>a, b</sup> | 1.24 | 0.36 | 0.70 | 2.19 | 0.463 |
| Model 3: *Fully Adjusted* <sup>c</sup> | 1.11 | 0.33 | 0.62 | 1.98 | 0.721 |

Notes: The *low-risk* group is the reference; RRR = relative risk ratio; SE = standard errors; CI = confidence interval; *p* = significance value.
a Demographic variables: age; sex
b Genetic variables: 10 principal components (PCs); C-reactive Protein (CRP) polygenic score (PGS); White Blood Cell Counts (WBCC) PGS; Insulin Growth Factor-1 (IGF-1) PGS; [Hair] Cortisol PGS; Sleep Duration PGS.
c All variables: age; sex; 10 PCs; CRP PGS; WBCC PGS; IGF-1 PGS; Cortisol PGS; education; wealth; occupational social status; smoking status; alcohol consumption; physical activity; mobility; limiting longstanding illness; health (i.e., chronic lung disease; coronary heart disease; abnormal heart rhythm; heart murmur; congestive heart failure; angina; hypertension; diabetes; cancer; Parkinson's; Alzheimer's; dementia; asthma; arthritis; osteoporosis; psychiatric disorder).

### Longitudinal associations of financial stress with suboptimal sleep

In models adjusted for baseline suboptimal sleep, age, sex, and genetic predisposition (Table 3), financial stress was longitudinally associated with a greater risk of experiencing short sleep (RRR=1.46; 95%CI=1.14-1.86; *p*=0.003), but not long sleep (RRR=1.22; 95%CI=0.86-1.72; *p*=0.265). When fully adjusted, financial stress was associated with a 31% greater risk of experiencing short sleep 4 years later (RRR=1.31; 95%CI=1.02-1.68; *p*=0.035).

**Table 3.**
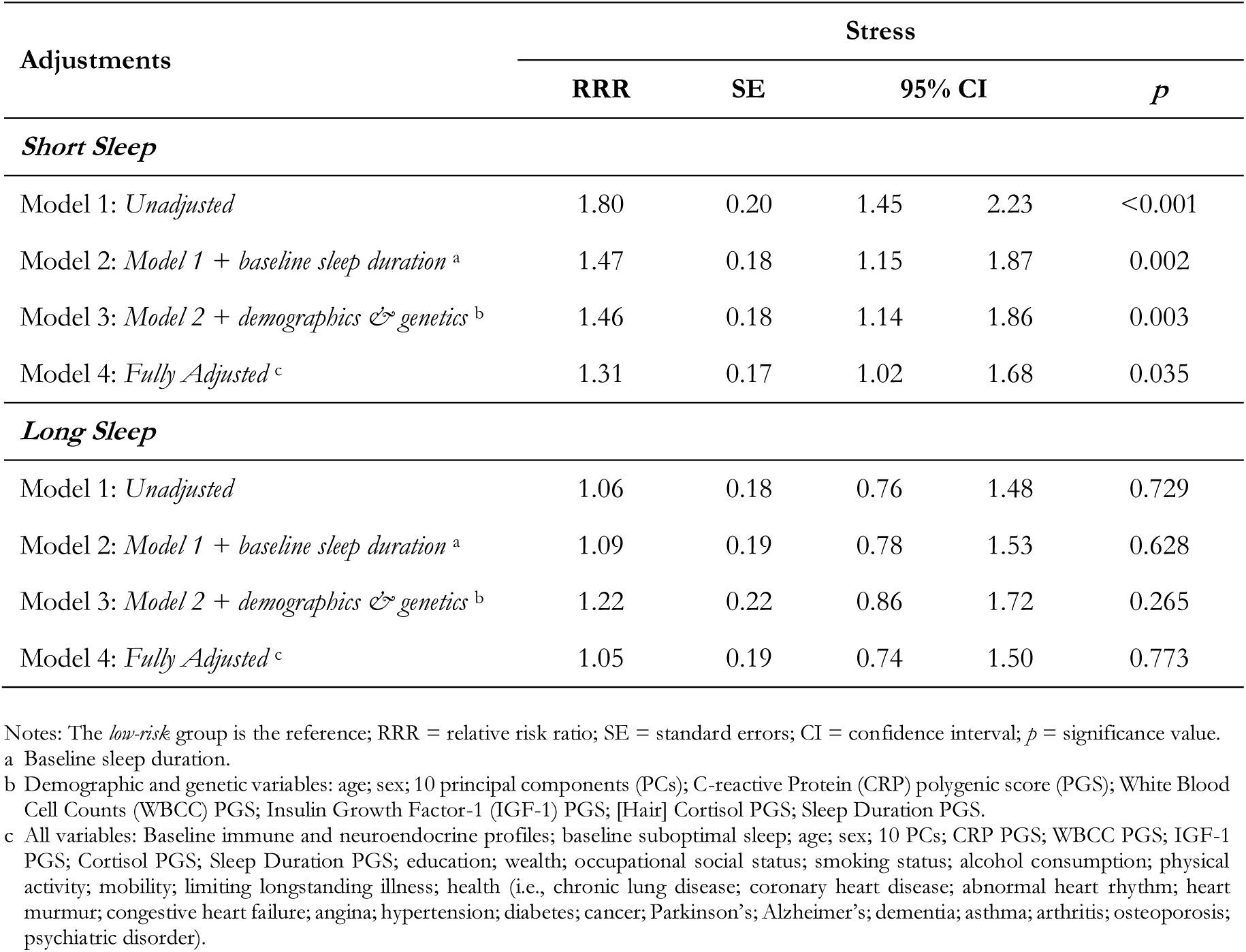
Longitudinal associations between stress and suboptimal sleep.

| Adjustments | Stress |  |  |  |  |
| --- | --- | --- | --- | --- | --- |
|  | RRR | SE | 95% CI |  | *p* |
| *Short Sleep* |  |  |  |  |  |
| Model 1: *Unadjusted* | 1.80 | 0.20 | 1.45 | 2.23 | <0.001 |
| Model 2: *Model 1 + baseline sleep duration* <sup>a</sup> | 1.47 | 0.18 | 1.15 | 1.87 | 0.002 |
| Model 3: *Model 2 + demographics & genetics* <sup>b</sup> | 1.46 | 0.18 | 1.14 | 1.86 | 0.003 |
| Model 4: *Fully Adjusted* <sup>c</sup> | 1.31 | 0.17 | 1.02 | 1.68 | 0.035 |
| *Long Sleep* |  |  |  |  |  |
| Model 1: *Unadjusted* | 1.06 | 0.18 | 0.76 | 1.48 | 0.729 |
| Model 2: *Model 1 + baseline sleep duration* <sup>a</sup> | 1.09 | 0.19 | 0.78 | 1.53 | 0.628 |
| Model 3: *Model 2 + demographics & genetics* <sup>b</sup> | 1.22 | 0.22 | 0.86 | 1.72 | 0.265 |
| Model 4: *Fully Adjusted* <sup>c</sup> | 1.05 | 0.19 | 0.74 | 1.50 | 0.773 |

Notes: The *low-risk* group is the reference; RRR = relative risk ratio; SE = standard errors; CI = confidence interval; *p* = significance value.
<sup>a</sup> Baseline sleep duration.
<sup>b</sup> Demographic and genetic variables: age; sex; 10 principal components (PCs); C-reactive Protein (CRP) polygenic score (PGS); White Blood Cell Counts (WBCC) PGS; Insulin Growth Factor-1 (IGF-1) PGS; [Hair] Cortisol PGS; Sleep Duration PGS.
<sup>c</sup> All variables: Baseline immune and neuroendocrine profiles; baseline suboptimal sleep; age; sex; 10 PCs; CRP PGS; WBCC PGS; IGF-1 PGS; Cortisol PGS; Sleep Duration PGS; education; wealth; occupational social status; smoking status; alcohol consumption; physical activity; mobility; limiting longstanding illness; health (i.e., chronic lung disease; coronary heart disease; abnormal heart rhythm; heart murmur; congestive heart failure; angina; hypertension; diabetes; cancer; Parkinson's; Alzheimer's; dementia; asthma; arthritis; osteoporosis; psychiatric disorder).

### Longitudinal associations of financial stress with immune and neuroendocrine profiles

Having adjusted for baseline profiles, along with demographic and genetic factors, as shown in Table 4, financial stress experience was longitudinally associated with 28% increased likelihood of being classified into the *moderate-risk* profile, as compared to the *low-risk* profile (RRR=1.28; 95%CI=1.06-1.54; *p*=0.010). But associations were reduced to null after full adjustment (RRR=1.18; 95%CI=0.97-1.43; *p*=0.093). By contrast, with membership of the *high-risk* profile across the same models, financial stress was associated with a 1.65 greater risk of belonging to this group (RRR=1.65; 95%CI=1.31-2.07; *p*<0.001), which fell to 1.42 after full adjustment (RRR=1.42; 95%CI=1.12-1.80; *p*=0.004).

**Table 4.**
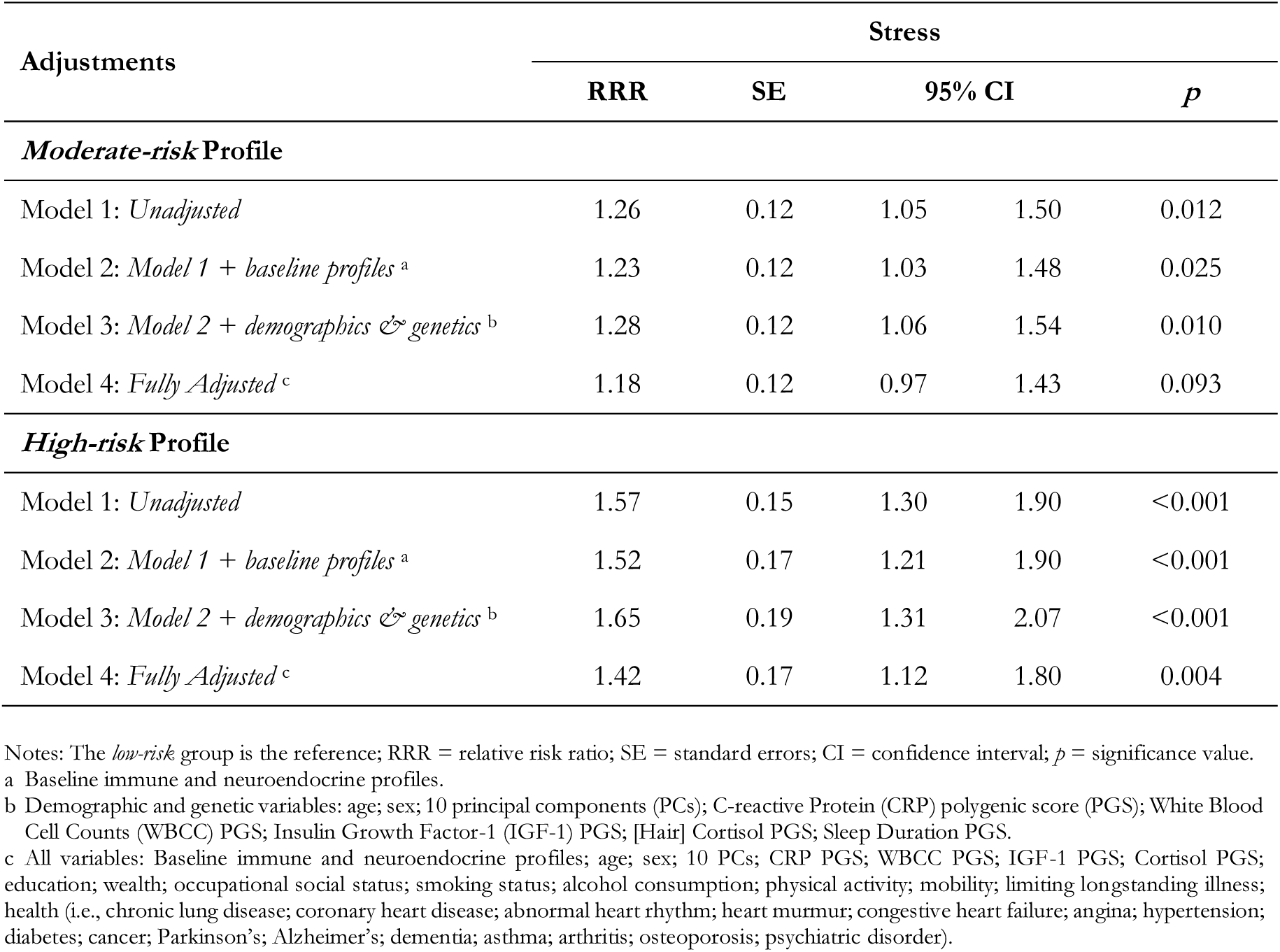
Longitudinal associations of stress with immune and neuroendocrine profiles.

### Longitudinal associations of suboptimal sleep with immune and neuroendocrine profiles

At any level of adjustment, suboptimal sleep durations were not prospectively associated with a greater risk of belonging to the *moderate-risk* profile compared with the *low-risk* profile (Table 5). And short sleep was not associated with future risk of *high-risk* immune-neuroendocrine profile membership in adjusted models. There were indications that long sleep was associated with a two-fold increase in the likelihood of *high-risk* profile membership when adjusted for baseline profiles, demographic, and genetic factors (RRR=2.02; 95%CI=1.02-3.98; p=0.043), but this association was attenuated to null after full adjustment (RRR=1.48; 95%CI=0.73-2.98; *p*=0.277).

**Table 5.**
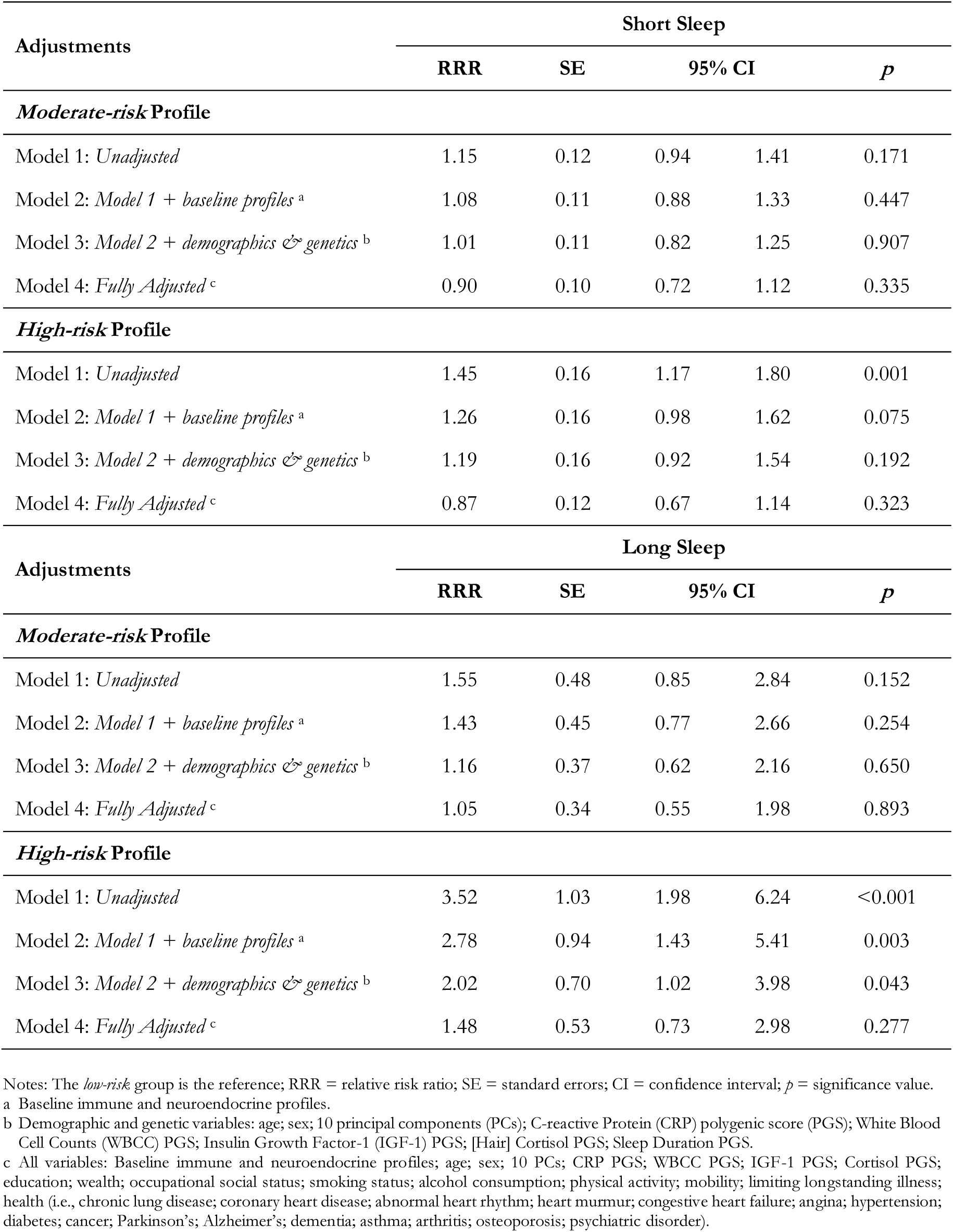
Longitudinal associations of suboptimal sleep with immune and neuroendocrine profiles.

### Effect modification between financial stress and suboptimal sleep in immune and neuroendocrine profile membership

There was no moderating effect of suboptimal sleep on the association between financial stress and immune-neuroendocrine profile membership (Table 6). The coefficients of interaction terms were close to zero for short sleep. The magnitude of effects were large for long sleep but with large confidence intervals. Thus, there was insufficient evidence to reject the null hypothesis.

**Table 6.**
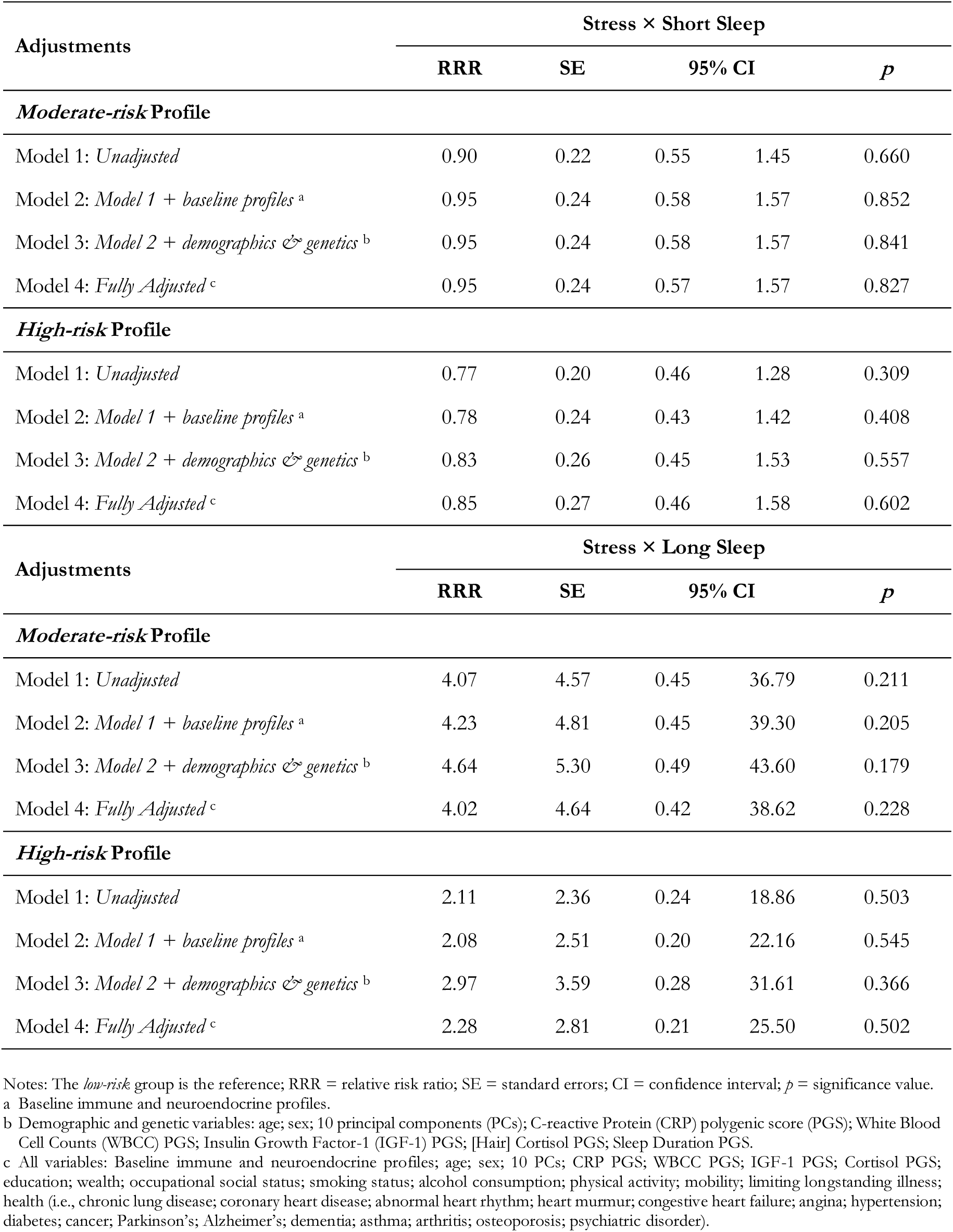
Effect modification between stress and suboptimal sleep in immune and neuroendocrine profile membership.

| Adjustments | Stress × Short Sleep |  |  |  |  |
| --- | --- | --- | --- | --- | --- |
|  | RRR | SE | 95% CI |  | *p* |
| ***Moderate-risk Profile*** |  |  |  |  |  |
| Model 1: *Unadjusted* | 0.90 | 0.22 | 0.55 | 1.45 | 0.660 |
| Model 2: *Model 1 + baseline profiles*<sup>a</sup> | 0.95 | 0.24 | 0.58 | 1.57 | 0.852 |
| Model 3: *Model 2 + demographics & genetics*<sup>b</sup> | 0.95 | 0.24 | 0.58 | 1.57 | 0.841 |
| Model 4: *Fully Adjusted*<sup>c</sup> | 0.95 | 0.24 | 0.57 | 1.57 | 0.827 |
| ***High-risk Profile*** |  |  |  |  |  |
| Model 1: *Unadjusted* | 0.77 | 0.20 | 0.46 | 1.28 | 0.309 |
| Model 2: *Model 1 + baseline profiles*<sup>a</sup> | 0.78 | 0.24 | 0.43 | 1.42 | 0.408 |
| Model 3: *Model 2 + demographics & genetics*<sup>b</sup> | 0.83 | 0.26 | 0.45 | 1.53 | 0.557 |
| Model 4: *Fully Adjusted*<sup>c</sup> | 0.85 | 0.27 | 0.46 | 1.58 | 0.602 |
| Adjustments | Stress × Long Sleep |  |  |  |  |
|  | RRR | SE | 95% CI |  | *p* |
| ***Moderate-risk Profile*** |  |  |  |  |  |
| Model 1: *Unadjusted* | 4.07 | 4.57 | 0.45 | 36.79 | 0.211 |
| Model 2: *Model 1 + baseline profiles*<sup>a</sup> | 4.23 | 4.81 | 0.45 | 39.30 | 0.205 |
| Model 3: *Model 2 + demographics & genetics*<sup>b</sup> | 4.64 | 5.30 | 0.49 | 43.60 | 0.179 |
| Model 4: *Fully Adjusted*<sup>c</sup> | 4.02 | 4.64 | 0.42 | 38.62 | 0.228 |
| ***High-risk Profile*** |  |  |  |  |  |
| Model 1: *Unadjusted* | 2.11 | 2.36 | 0.24 | 18.86 | 0.503 |
| Model 2: *Model 1 + baseline profiles*<sup>a</sup> | 2.08 | 2.51 | 0.20 | 22.16 | 0.545 |
| Model 3: *Model 2 + demographics & genetics*<sup>b</sup> | 2.97 | 3.59 | 0.28 | 31.61 | 0.366 |
| Model 4: *Fully Adjusted*<sup>c</sup> | 2.28 | 2.81 | 0.21 | 25.50 | 0.502 |

Notes: The *low-risk* group is the reference; RRR = relative risk ratio; SE = standard errors; CI = confidence interval; *p* = significance value.
<sup>a</sup> Baseline immune and neuroendocrine profiles.
<sup>b</sup> Demographic and genetic variables: age; sex; 10 principal components (PCs); C-reactive Protein (CRP) polygenic score (PGS); White Blood Cell Counts (WBCC) PGS; Insulin Growth Factor-1 (IGF-1) PGS; [Hair] Cortisol PGS; Sleep Duration PGS.
<sup>c</sup> All variables: Baseline immune and neuroendocrine profiles; age; sex; 10 PCs; CRP PGS; WBCC PGS; IGF-1 PGS; Cortisol PGS; education; wealth; occupational social status; smoking status; alcohol consumption; physical activity; mobility; limiting longstanding illness; health (i.e., chronic lung disease; coronary heart disease; abnormal heart rhythm; heart murmur; congestive heart failure; angina; hypertension; diabetes; cancer; Parkinson's; Alzheimer's; dementia; asthma; arthritis; osteoporosis; psychiatric disorder).

**Table 7.** Longitudinal associations of immune and neuroendocrine profiles with suboptimal sleep.

| Adjustments | Moderate-risk Profile |  |  |  |  |
| --- | --- | --- | --- | --- | --- |
|  | RRR | SE | 95% CI |  | *p* |
| Short Sleep |  |  |  |  |  |
| Model 1: *Unadjusted* | 1.51 | 0.15 | 1.24 | 1.84 | <0.001 |
| Model 2: *Model 1 + baseline profiles*<sup>a</sup> | 1.38 | 0.15 | 1.11 | 1.71 | 0.003 |
| Model 3: *Model 2 + demographics & genetics*<sup>b</sup> | 1.38 | 0.15 | 1.11 | 1.72 | 0.004 |
| Model 4: *Fully Adjusted*<sup>c</sup> | 1.21 | 0.14 | 0.96 | 1.51 | 0.103 |
| Adjustments | High-risk Profile |  |  |  |  |
|  | RRR | SE | 95% CI |  | *p* |
| Model 1: *Unadjusted* | 1.33 | 0.20 | 0.99 | 1.79 | 0.059 |
| Model 2: *Model 1 + baseline profiles*<sup>a</sup> | 1.13 | 0.19 | 0.82 | 1.57 | 0.454 |
| Model 3: *Model 2 + demographics & genetics*<sup>b</sup> | 1.11 | 0.19 | 0.80 | 1.54 | 0.539 |
| Model 4: *Fully Adjusted*<sup>c</sup> | 0.86 | 0.15 | 0.61 | 1.22 | 0.388 |
| Adjustments | Moderate-risk Profile |  |  |  |  |
|  | RRR | SE | 95% CI |  | *p* |
| Long Sleep |  |  |  |  |  |
| Model 1: *Unadjusted* | 2.14 | 0.31 | 1.61 | 2.83 | <0.001 |
| Model 2: *Model 1 + baseline profiles*<sup>a</sup> | 2.11 | 0.31 | 1.58 | 2.80 | <0.001 |
| Model 3: *Model 2 + demographics & genetics*<sup>b</sup> | 1.91 | 0.28 | 1.43 | 2.56 | <0.001 |
| Model 4: *Fully Adjusted*<sup>c</sup> | 1.66 | 0.25 | 1.23 | 2.23 | 0.001 |
| Adjustments | High-risk Profile |  |  |  |  |
|  | RRR | SE | 95% CI |  | *p* |
| Model 1: *Unadjusted* | 2.93 | 0.53 | 2.06 | 4.18 | <0.001 |
| Model 2: *Model 1 + baseline profiles*<sup>a</sup> | 2.81 | 0.52 | 1.96 | 4.03 | <0.001 |
| Model 3: *Model 2 + demographics & genetics*<sup>b</sup> | 2.30 | 0.43 | 1.59 | 3.33 | <0.001 |
| Model 4: *Fully Adjusted*<sup>c</sup> | 1.81 | 0.35 | 1.24 | 2.64 | 0.002 |

Notes: The *low-risk* group is the reference; RRR = relative risk ratio; SE = standard errors; CI = confidence interval; *p* = significance value.
<sup>a</sup> Baseline immune and neuroendocrine profiles.
<sup>b</sup> Demographic and genetic variables: age; sex; 10 principal components (PCs); C-reactive Protein (CRP) polygenic score (PGS); White Blood Cell Counts (WBCC) PGS; Insulin Growth Factor-1 (IGF-1) PGS; [Hair] Cortisol PGS; Sleep Duration PGS.
<sup>c</sup> All variables: Baseline immune and neuroendocrine profiles; age; sex; 10 PCs; CRP PGS; WBCC PGS; IGF-1 PGS; Cortisol PGS; education; wealth; occupational social status; smoking status; alcohol consumption; physical activity; mobility; limiting longstanding illness; health (i.e., chronic lung disease; coronary heart disease; abnormal heart rhythm; heart murmur; congestive heart failure; angina; hypertension; diabetes; cancer; Parkinson's; Alzheimer's; dementia; asthma; arthritis; osteoporosis; psychiatric disorder).

### Sensitivity Analyses

First, when associations were stratified by age, financial stress was associated with a 70% higher relative risk of belonging to the *high-risk* immune-neuroendocrine profile in younger individuals (RRR=1.70; 95%CI=1.20-2.41; *p*=0.003), with indications of a gradient in risk, such that the *moderate-risk* profile showed an intermediate pattern (RRR=1.30; 95%CI=1.00-1.70; *p*=0.050). However, there were no associations in the older age category (Table S4). As it relates to the suboptimal sleep associations with immune-neuroendocrine profile membership, effect sizes did not differ by age (Table S5). In the second sensitivity analysis, sex-stratified analyses revealed that men, but not women, stressed from a lack of resources had a greater risk of belonging to the *high-risk* immune-neuroendocrine profile (RRR=1.68; 95%CI=1.16-2.43; *p*=0.006; Table S6) despite an overlap of confidence intervals. But this pattern between the sexes was not replicated in suboptimal sleep associations with immune-neuroendocrine profile membership (Table S7). The third sensitivity analysis included BMI as an additional covariate in Model 4, with no substantive change seen in the magnitude of effects (Table S9). The fourth sensitivity analysis used less stringent thresholds for short and long sleep (≤6 hr [*n*=2,095; 19.29%]; ≥8 hr [*n*=4,788; 44.08%] respectively) and results were materially unchanged (Table S10). In the final sensitivity analysis, a 1± increase in genetic liability for short sleep and long was not associated with immune-neuroendocrine profile membership (Table S8).

## Discussion

In a large, population-representative, prospective cohort study of older adults, cross-sectional, longitudinal, and multiplicative associations were tested between financial stress and suboptimal sleep in immune-neuroendocrine profile membership. Financial stress was cross-sectionally and longitudinally associated with short sleep, but not long sleep. Associations held having controlled for genetic predisposition and a broad selection of covariates. Financial stress was additionally associated with *high-risk* profile membership four years later. It is worth noting that there was no change in the magnitude of effects when accounting for BMI. In addition, against our expectations, suboptimal sleep was not associated with immune-neuroendocrine profile membership, nor did it modify the relationship between financial stress and said profiles.

The preliminary findings extend a growing evidence-base that stress exposure has a significant liability on sleep routines (Beck et al., 2022; Slavish et al., 2021; Yap et al., 2020), credibly through the activation the HPA- and SNS-axes (Kalmbach et al., 2018). But here we add more specificity, with financial stress being a more salient risk factor to short sleep than long sleep. This is persuasive given that different molecular mechanisms are thought to underlie associations at the either end of the sleep duration distribution (Hamilton, Steptoe, et al., 2023). Contrasts with other studies are likely due to differences in sleep definitions, objectivity, and architecture, together with the acuteness and nature of stress (S. Y. Kim et al., 2020; Vandekerckhove et al., 2011; Zagaria et al., 2023).

Our analytical strategy leveraged much more than a convenient device with which to model individual differences. Semiparametric, finite mixture models, such as the LPA used here, offer a principled way to identify heterogeneity based on observed patterns within empirical data (S. H. Kim & Mokhtarian, 2023). It proves useful when the normality assumption of the maximum likelihood (ML) fitting function, used to estimate the model, is unwittingly violated (Cole & Bauer, 2016). These models avoid the need for *ad hoc* classifications. Instead they are estimated in the service of more flexibly modelling the characteristics of the aggregate population as a whole (Bauer, 2005). While studies that isolate individual biomarkers offer detailed mechanistic insights, studies that aggregate profiles contribute to a more comprehensive understanding of system-level activity. In the present study, distinct biological groups in the population were identified. Where financial stress was an antecedent, differences in the risk of belonging to the *moderate*- or *high-risk* groups, as compared to the *low-risk* group, emerged. This emphasis on group-level differences highlights the importance of considering the biological context and complexity. And results advance a precision medicine approach, where remedies can be directed toward identified subgroups, rather than indiscriminate treatments across heterogeneous populations.

Beyond a binary interpretation of statistical significance, the interpretation of estimates must consider whether the effect size is meaningful from a public health perspective. An evaluation that is contingent on exposure and outcome prevalence, together with the Minimal Clinically Important Difference (MCID), which captures the magnitude of the issue and its ecological value (McGlothlin & Lewis, 2014). Given the ubiquity of financial stress exposure and the far-reaching impacts that biological processes have on health, a 42% increase in relative risk is meaningful. While there are notable complications to drawing policy conclusions from a single study (Bann et al., 2024), financial stress associations with immune-neuroendocrine profile memberships echoes earlier work in the same longitudinal dataset (Hamilton, Iob, et al., 2023). This result can, therefore, be treated as scientific replication that underscores the robustness and consistency of the observed relationships, despite a more comprehensive biomarker matrix included into the LPA here and more exhaustive controls used to mitigate confounding.

The unexpected finding that suboptimal sleep was not prospectively associated with immune-neuroendocrine profile membership is an important contribution to understanding the role of sleep in immune-neuroendocrine processes. It challenges compelling evidence elsewhere that sleep is integral to endocrine, metabolic, and immune integrity, and where sleep loss, even acutely, perturbs gene expression and the functional cellular response (Garbarino et al., 2021; Irwin, 2006). Meta-analytic evidence substantiates cross-sectional associations between suboptimal sleep and heightened inflammatory levels (Irwin et al., 2016), with further evidence that associations are longitudinal (Cho et al., 2015; Ferrie et al., 2013). Even when modelled curvilinearly, sleep duration remained associated with inflammatory processes (Prather et al., 2015). But in all cases, results depended on the biomarkers measured and the temporal relationship between exposures and outcomes.

Sleep duration likely activates pro-inflammatory pathways (Patel et al., 2009). But one plausible explanation for the contrasting findings in the present study is that suboptimal sleep may exert its effects on specific immune, neural, and endocrine biomarkers (Ferrie et al., 2013; Irwin et al., 2016; Prather et al., 2015). And it may do this without contemporaneously influencing a constellation of biomarkers, as would be captured by the latent profiles that represent co-variation across multiple biomarkers at a single timepoint (Morin et al., 2016). In this way, the profiles would not be as sensitive to suboptimal sleep durations as isolated biomarkers, because they represent greater physiological stability, following a more adaptive response to exogenous-endogenous challenge, through systemic-level compensatory biological mechanisms that dilute exposure impact.

Polygenic risk prediction is a useful tool in observational studies to confirm the less confounded role of traits in outcomes, albeit limited to genetic contribution (Hamilton, Steptoe, et al., 2023). Results arising from the PGSs for short and long sleep, support the phenotypic findings that suboptimal sleep durations are not prospectively associated with immune-neuroendocrine profiles. These corresponding results raise the possibility of reverse causality. First, having considered the previously reported role of immune-neuroendocrine biomarkers in diurnal variations (Besedovsky et al., 2019; Irwin, 2019), with much of the evidence on sleep duration and inflammation being cross-sectional, limiting inferences on directionality (Irwin et al., 2016; Patel et al., 2009). Second, because PGSs point to directionality given they are largely unconfounded; predicated on inherited DNA differences from birth that increase the predictive facility of complex traits in unrelated individuals among the population (Plomin & Von Stumm, 2022; Wray et al., 2021). Still, the current predictive facility of PGSs is insufficient for clinical implementation. This is due to the aetiological complexity of common disorders, like sleep duration, that are influenced by gene-environment interactions, with polygenic architecture that has contributions from multiple single nucleotide polymorphism (SNPs) of small effect, rather than a dominant genetic variant of large effect that can be isolated (Pain et al., 2021). Owing to this complexity, the ecological value of PGSs is reduced and the immediate scope to dismiss the credibility of sleep-related policy from these findings is narrow. Thus, results can be considered preliminary and hypothesis-generating.

It is, nonetheless, important to control for genetic influence in analyses, given the stability of genetic sequence across the lifespan and genetic variation that accounts for a notable proportion of individual difference in health and disease (Polderman et al., 2015). Our adjustment for a comprehensive selection of confounders, notably genetic predisposition for adverse biomarker levels, has rarely been reported elsewhere. We show that the longitudinal role of financial stress on suboptimal sleep and immune-neuroendocrine profiles is independent of genetic predisposition. But the role of suboptimal sleep on profile membership was not robust enough to withstand this genetic influence. The contrast with earlier evidence linking suboptimal sleep to inflammatory markers, may, in part, be attributable to statistically less controlled analyses that increase the likelihood of Type I error (Strain et al., 2020).

The age composition of the cohort has important implications. There is suggestive evidence of age-related effects from the first sensitivity analyses that warrants replication in a younger sample. One notion that merits consideration is survivor bias (viz. left truncation), where selective attrition led to a healthier, more resilient older group upon stratification, who are not characteristic of the general population (Vansteelandt et al., 2018). Moreover, the practice of napping might serve as a protective factor for immune-neuroendocrine processes. One study found cortisol decreased immediately after a midday nap (limited to 30 minutes), and this was accompanied by a return to baseline leukocytes counts (Faraut et al., 2011). In a later randomised, polysomnography-monitored study, an increase in norepinephrine and cytokine values after a sleep-restricted night was not observed after the countermeasure of a nap (Faraut et al., 2015). Given the higher prevalence of napping among older cohorts (Kocevska et al., 2020), this practice may have mitigated the influence of sleep duration on the inflammatory status of this population by acting on homeostatic processes. Equally, this cohort may not expend as much energy as younger cohorts (Gomes et al., 2017), with insufficient sleep debt corresponding to lesser homeostatic pressure (i.e., compensatory increases in essential sleep duration, sleep consolidation, and sleep intensity that mounts in response to an extended period of wakefulness (Dittrich et al., 2015). This pressure accumulates exponentially during wakefulness and dissipates during sleep, striking balance between sleep-promoting and wake-promoting neurons that control the volume and amplitude of slow-wave activity (Taillard et al., 2021).

In the absence of moderation, results putatively contradict anecdotal and empirical evidence (Chiang et al., 2019) that stress exposure compounded with suboptimal sleep durations increase risk to biological signatures. Analyses can be considered preliminary, owing to the modest cell sizes of suboptimal sleep cases, and other common power attenuating factors, including exposure-mediator intercorrelations, scale coarseness, the artificial categorisations of continuous variables, or the transformation of non-normal outcomes (Dawson, 2014). These may have restricted the detection of interactions, with unstable parameter estimates and inflated standard errors, but given the sizeable alphas, it is unlikely that a greater powered sample would have detected moderating effects with reliable estimates. Results point to financial stress being a more proximate risk factor to immune-neuroendocrine processes than suboptimal sleep, so much so that financial stress may reach a threshold beyond which suboptimal sleep has limited additional returns. The lack of interaction is consistent with the limited influence of sleep duration in this cohort (Taillard et al., 2021).

Invariably, findings must be interpreted in light of some limitations. Financial stress and sleep duration are self-reported and time varying, so individuals may experience changes throughout the course of the study that are subject to recall bias. Future study may benefit from objective measures in a time-varying effect model (Tan et al., 2012). Moreover, financial stress was measured with a single item, so it may not have captured the multidimensionality of material deprivation (Hamilton & Steptoe, 2022). While the causal DAG represented a particular set of assumptions, its complexity does not explicitly reflect real-world concerns about reciprocity or sources of bias, and it does not specify the estimate magnitude, nor its interplay with random errors (Lipsky & Greenland, 2022). Observational studies cannot yield definitive conclusions on cause and effect, particularly in the face of reverse causality (Sattar & Preiss, 2017). In this respect, while not without limitations, Mendelian randomization (MR) offers a promising avenue to overcome this challenge. But the most promising approach is to triangulate evidence across multiple sources to improve causal claims (Lawlor et al., 2017).

## Conclusion

Immune and neuroendocrine activity has a highly complex aetiology that arises from multifaceted interplays across biological, physiological, behavioural, social, and environmental dynamics. Capitalising on converging methods of estimation, we unravel a portion of this complexity, by giving insight into the independent and interactive roles of financial stress and suboptimal sleep in immune-neuroendocrine latent profile membership. Financial-related stress remains a reliable target for reducing short-sleep and offers a promising pathway for understanding the stratification of immune-neuroendocrine activity among the general population. Policies to screen for and alleviate the financial burden on the population may be an effective strategy to reduce adverse immune-endocrine processes that lead to generalised disease, with a view to mitigate the liability on an already stretched public health system. While suboptimal sleep has long been recognised as a primal factor in biological processes, its inconsistent findings make it a less compelling target for immune-endocrine patterning, certainly in older adults. And there was no conclusive evidence to support that the combined experience of financial stress and suboptimal sleep is worse for immune-neuroendocrine patterning than financial stress alone in this group. This study underscores several potential areas of interest for future research while emphasising the need for translational investigations. Most pressing among them is the need to determine whether results hold in a younger cohort, whether associations are causal, and whether these immune-neuroendocrine profiles mediate associations between stress and disease. Addressing these would contribute to the understanding of biosocial mechanisms to disease.

## Supporting information

Full Supplement

## Data Availability

All data produced in the present study are available upon reasonable request to the authors.

## Data Sharing

The data are deposited in the UK Data Archive and freely available through the UK Data Service (SN 8688 and 5050) and can be accessed here: discover.ukdataservice.ac.uk.

## Ethical Approval

The National Research Ethics Service (London Multicentre Research Ethics Committee [MREC/01/2/91] nres.npsa.nhs.uk) granted ethical approval for each of the ELSA waves. All participants provided informed consent, and research was performed in accordance with research and data protection guidelines.

## Contributorship

Study funding was secured by AS. Conception and planning by both authors. OSH designed the statistical analysis plan, prepared the data, performed the analyses, and drafted the manuscript. Both authors interpreted results and act as guarantors, taking responsibility for integrity of the data, the accuracy of analyses, its interpretation, and the critical appraisal the manuscript.

---

**Figure 1.**
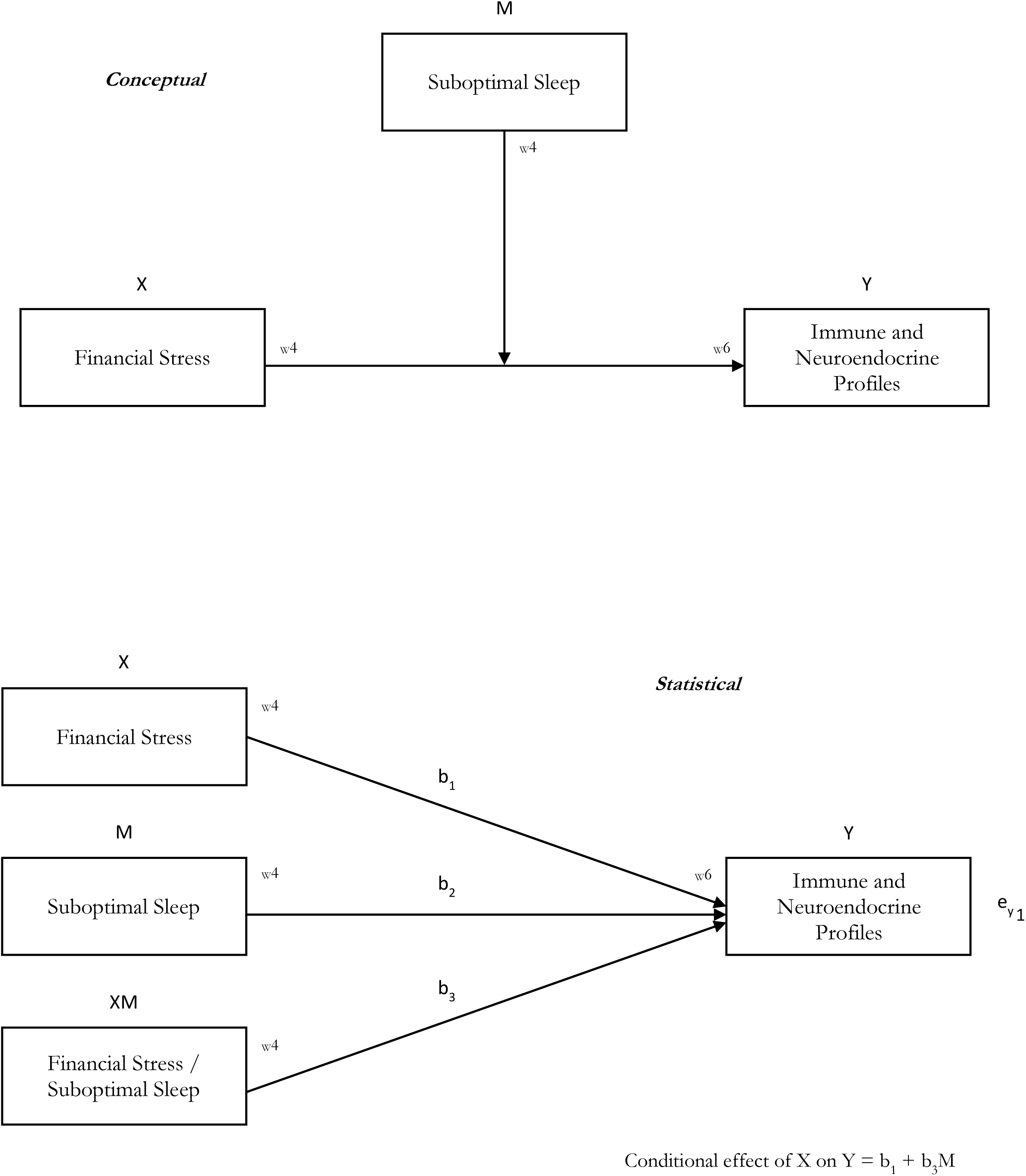
A Conceptual and Statistical Diagram of Effect Modification.

**Figure 2.**
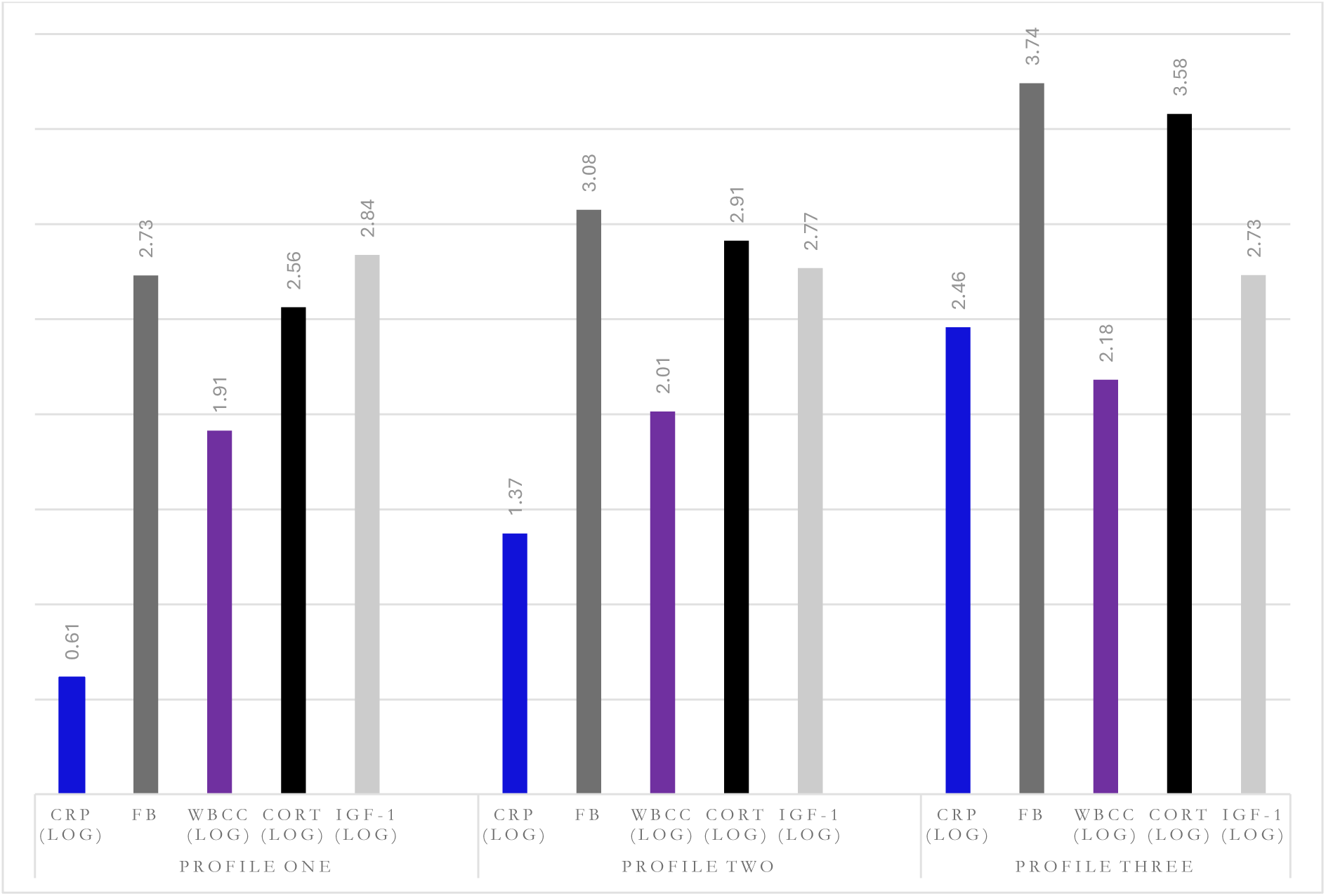
Predicted Marginal Mean Margins of Immune and Neuroendocrine Profiles.

## REFERENCES

Aguinis, H., & Gottfredson, R. K. (2010). Best-practice recommendations for estimating interaction effects using moderated multiple regression.

Bakour, C., Schwartz, S., O’Rourke, K., Wang, W., Sappenfield, W., Couluris, M., & Chen, H. (2017). Sleep Duration Trajectories and Systemic Inflammation in Young Adults: Results From the National Longitudinal Study of Adolescent to Adult Health (Add Health). Sleep, 40(11). 10.1093/sleep/zsx156

Bale, T. L., & Epperson, C. N. (2015). Sex differences and stress across the lifespan. Nature Neuroscience, 18(10), 1413–1420. 10.1038/nn.4112

Bann, D., Courtin, E., Davies, N. M., & Wright, L. (2024). Dialling back ‘impact’ claims: Researchers should not be compelled to make policy claims based on single studies. International Journal of Epidemiology, 53(1), dyad181. 10.1093/ije/dyad181

Bauer, D. J. (2005). A Semiparametric Approach to Modeling Nonlinear Relations Among Latent Variables. Structural Equation Modeling: A Multidisciplinary Journal, 12(4), 513–535. 10.1207/s15328007sem1204_1

Becher, B., Spath, S., & Goverman, J. (2017). Cytokine networks in neuroinflammation. Nature Reviews Immunology, 17(1), 49–59. 10.1038/nri.2016.123

Beck, J., Loretz, E., & Rasch, B. (2022). Stress dynamically reduces sleep depth: Temporal proximity to the stressor is crucial. Cerebral Cortex, 33(1), 96–113. 10.1093/cercor/bhac055

Besedovsky, L., Lange, T., & Haack, M. (2019). The Sleep-Immune Crosstalk in Health and Disease. Physiological Reviews, 99(3), 1325–1380. 10.1152/physrev.00010.2018

Bierhaus, A., Humpert, P. M., & Nawroth, P. P. (2006). Linking Stress to Inflammation. Anesthesiology Clinics of North America, 24(2), 325–340. 10.1016/j.atc.2006.01.001

Brydon, L., Wright, C. E., O’Donnell, K., Zachary, I., Wardle, J., & Steptoe, A. (2008). Stress-induced cytokine responses and central adiposity in young women. International Journal of Obesity, 32(3), 443–450. 10.1038/sj.ijo.0803767

Chiang, J. J., Cole, S. W., Bower, J. E., Irwin, M. R., Taylor, S. E., Arevalo, J., & Fuligni, A. J. (2019). Daily interpersonal stress, sleep duration, and gene regulation during late adolescence. Psychoneuroendocrinology, 103, 147–155. 10.1016/j.psyneuen.2018.11.026

Cho, H. J., Seeman, T. E., Kiefe, C. I., Lauderdale, D. S., & Irwin, M. R. (2015). Sleep disturbance and longitudinal risk of inflammation: Moderating influences of social integration and social isolation in the Coronary Artery Risk Development in Young Adults (CARDIA) study. *Brain*, Behavior, and Immunity, 46, 319–326. 10.1016/j.bbi.2015.02.023

Cole, V. T., & Bauer, D. J. (2016). A Note on the Use of Mixture Models for Individual Prediction. Structural Equation Modeling: A Multidisciplinary Journal, 23(4), 615–631. 10.1080/10705511.2016.1168266

Dawson, J. F. (2014). Moderation in Management Research: What, Why, When, and How. Journal of Business and Psychology, 29(1), 1–19. 10.1007/s10869-013-9308-7

Dittrich, L., Morairty, S. R., Warrier, D. R., & Kilduff, T. S. (2015). Homeostatic Sleep Pressure is the Primary Factor for Activation of Cortical nNOS/NK1 Neurons. Neuropsychopharmacology, 40(3), 632–639. 10.1038/npp.2014.212

Faraut, B., Boudjeltia, K. Z., Dyzma, M., Rousseau, A., David, E., Stenuit, P., Franck, T., Antwerpen, P. V., Vanhaeverbeek, M., & Kerkhofs, M. (2011). Benefits of napping and an extended duration of recovery sleep on alertness and immune cells after acute sleep restriction☆. Brain, Behavior, and Immunity, 25(1), 16–24. 10.1016/j.bbi.2010.08.001

Faraut, B., Nakib, S., Drogou, C., Elbaz, M., Sauvet, F., De Bandt, J.-P., & Léger, D. (2015). Napping Reverses the Salivary Interleukin-6 and Urinary Norepinephrine Changes Induced by Sleep Restriction. The Journal of Clinical Endocrinology & Metabolism, 100(3), E416–E426. 10.1210/jc.2014-2566

Ferrie, J. E., Kivimaki, M., Akbaraly, T. N., Singh-Manoux, A., Miller, M. A., Gimeno, D., Kumari, M., Davey Smith, G., & Shipley, M. J. (2013). Associations Between Change in Sleep Duration and Inflammation: Findings on C-reactive Protein and Interleukin 6 in the Whitehall II Study. American Journal of Epidemiology, 178(6), 956–961. 10.1093/aje/kwt072

Furman, D., Campisi, J., Verdin, E., Carrera-Bastos, P., Targ, S., Franceschi, C., Ferrucci, L., Gilroy, D. W., Fasano, A., Miller, G. W., Miller, A. H., Mantovani, A., Weyand, C. M., Barzilai, N., Goronzy, J. J., Rando, T. A., Effros, R. B., Lucia, A., Kleinstreuer, N., & Slavich, G. M. (2019). Chronic inflammation in the etiology of disease across the life span. Nature Medicine, 25(12), 1822–1832. 10.1038/s41591-019-0675-0

Garbarino, S., Lanteri, P., Bragazzi, N. L., Magnavita, N., & Scoditti, E. (2021). Role of sleep deprivation in immune-related disease risk and outcomes. Communications Biology, 4(1), 1304. 10.1038/s42003-021-02825-4

Garfield, V., Fatemifar, G., Dale, C., Smart, M., Bao, Y., Llewellyn, C. H., Steptoe, A., Zabaneh, D., & Kumari, M. (2019). Assessing potential shared genetic aetiology between body mass index and sleep duration in 142,209 individuals. Genetic Epidemiology, 43(2), 207–214. 10.1002/gepi.22174

Gomes, M., Figueiredo, D., Teixeira, L., Poveda, V., Paúl, C., Santos-Silva, A., & Costa, E. (2017). Physical inactivity among older adults across Europe based on the SHARE database. Age and Ageing, 46(1), 71–77. 10.1093/ageing/afw165

Graham, J. E., Christian, L. M., & Kiecolt-Glaser, J. K. (2006). Stress, Age, and Immune Function: Toward a Lifespan Approach | Journal of Behavioral Medicine. https://link.springer.com/article/10.1007/s10865-006-9057-4#citeas

Hainmueller, J., Mummolo, J., & Xu, Y. (2018). How Much Should We Trust Estimates from Multiplicative Interaction Models? Simple Tools to Improve Empirical Practice. Political Analysis.

Hamilton, O. S., Iob, E., Ajnakina, O., Kirkbride, J. B., & Steptoe, A. (2023). Immune-neuroendocrine patterning and response to stress. A latent profile analysis in the English longitudinal study of ageing. Brain, Behavior, and Immunity, S0889159123003458. 10.1016/j.bbi.2023.11.012

Hamilton, O. S., & Steptoe, A. (2022). Socioeconomic determinants of inflammation and neuroendocrine activity: A longitudinal analysis of compositional and contextual effects. Brain, Behavior, and Immunity, 107, 276–285. 10.1016/j.bbi.2022.10.010

Hamilton, O. S., Steptoe, A., & Ajnakina, O. (2023). Polygenic predisposition, sleep duration, and depression: Evidence from a prospective population-based cohort. Translational Psychiatry, 13(1), 323. 10.1038/s41398-023-02622-z

Hernán, M. A., & Monge, S. (2023). Selection bias due to conditioning on a collider. BMJ, p1135. 10.1136/bmj.p1135

Irwin, M. R. (2006). Sleep Deprivation and Activation of Morning Levels of Cellular and Genomic Markers of Inflammation. Archives of Internal Medicine, 166(16), 1756. 10.1001/archinte.166.16.1756

Irwin, M. R. (2019). Sleep and inflammation: Partners in sickness and in health. Nature Reviews Immunology, 19(11), Article 11. 10.1038/s41577-019-0190-z

Irwin, M. R., Olmstead, R., & Carroll, J. E. (2016). Sleep Disturbance, Sleep Duration, and Inflammation: A Systematic Review and Meta-Analysis of Cohort Studies and Experimental Sleep Deprivation. Biological Psychiatry, 80(1), 40–52. 10.1016/j.biopsych.2015.05.014

Jerison, E. (2024). Dynamical control of immunity and inflammation. Biophysical Journal, 123(3), 309a. 10.1016/j.bpj.2023.11.1908

Jones, C., & Gatchel, R. J. (2018). Palliative Medicine: The Importance of Sleep, Stress, and Behavior. Palliative Medicine & Care Open Access, 5(2). https://symbiosisonlinepublishing.com/palliative-medicine-care/palliative-medicine-care58.php

Kalmbach, D., Cuamatzi-Castelan, A., Tonnu, C., Tran, K. M., Anderson, J., Roth, T., & Drake, C. (2018). Hyperarousal and sleep reactivity in insomnia: Current insights. *Nature and Science of Sleep*, Volume 10, 193–201. 10.2147/NSS.S138823

Kim, S. H., & Mokhtarian, P. L. (2023). Finite mixture (or latent class) modeling in transportation: Trends, usage, potential, and future directions. Transportation Research Part B: Methodological, 172, 134–173. 10.1016/j.trb.2023.03.001

Kim, S. Y., Kark, S. M., Daley, R. T., Alger, S. E., Rebouças, D., Kensinger, E. A., & Payne, J. D. (2020). Interactive effects of stress reactivity and rapid eye movement sleep theta activity on emotional memory formation. Hippocampus, 30(8), 829–841. 10.1002/hipo.23138

Kocevska, D., Lysen, T. S., Dotinga, A., Koopman-Verhoeff, M. E., Luijk, M. P. C. M., Antypa, N., Biermasz, N. R., Blokstra, A., Brug, J., Burk, W. J., Comijs, H. C., Corpeleijn, E., Dashti, H. S., De Bruin, E. J., De Graaf, R., Derks, I. P. M., Dewald-Kaufmann, J. F., Elders, P. J. M., Gemke, R. J. B. J., … Tiemeier, H. (2020). Sleep characteristics across the lifespan in 1.1 million people from the Netherlands, United Kingdom and United States: A systematic review and meta-analysis. Nature Human Behaviour, 5(1), 113–122. 10.1038/s41562-020-00965-x

Lawlor, D. A., Tilling, K., & Davey Smith, G. (2017). Triangulation in aetiological epidemiology. International Journal of Epidemiology, 45(6), 1866–1886. 10.1093/ije/dyw314

Lipsky, A. M., & Greenland, S. (2022). Causal Directed Acyclic Graphs. JAMA, 327(11), 1083. 10.1001/jama.2022.1816

McEwen, B. S. (2007). Physiology and Neurobiology of Stress and Adaptation: Central Role of the Brain. Physiological Reviews, 87(3), 873–904. 10.1152/physrev.00041.2006

McGlothlin, A. E., & Lewis, R. J. (2014). Minimal Clinically Important Difference: Defining What Really Matters to Patients. JAMA, 312(13), 1342. 10.1001/jama.2014.13128

Minkel, J., Moreta, M., Muto, J., Htaik, O., Jones, C., Basner, M., & Dinges, D. (2014). Sleep deprivation potentiates HPA axis stress reactivity in healthy adults. *Health Psychology: Official Journal of the Division of Health Psychology*, American Psychological Association, 33(11), 1430–1434. 10.1037/a0034219

Morin, A. J. S., Meyer, J. P., Creusier, J., & Biétry, F. (2016). Multiple-Group Analysis of Similarity in Latent Profile Solutions. Organizational Research Methods, 19(2), 231–254. 10.1177/1094428115621148

Muscatell, K. A., Brosso, S. N., & Humphreys, K. L. (2020). Socioeconomic status and inflammation: A meta-analysis. Molecular Psychiatry, 25(9), Article 9. 10.1038/s41380-018-0259-2

Pain, O., Gillett, A. C., Austin, J. C., Folkersen, L., & Lewis, C. M. (2022). A tool for translating polygenic scores onto the absolute scale using summary statistics. European Journal of Human Genetics, 30(3), 339–348. 10.1038/s41431-021-01028-z

Pain, O., Glanville, K. P., Hagenaars, S. P., Selzam, S., Fürtjes, A. E., Gaspar, H. A., Coleman, J. R. I., Rimfeld, K., Breen, G., Plomin, R., Folkersen, L., & Lewis, C. M. (2021). Evaluation of polygenic prediction methodology within a reference-standardized framework. PLOS Genetics, 17(5), e1009021. 10.1371/journal.pgen.1009021

Patel, S. R., Zhu, X., Storfer-Isser, A., Mehra, R., Jenny, N. S., Tracy, R., & Redline, S. (2009). Sleep Duration and Biomarkers of Inflammation. Sleep, 32(2), 200–204. 10.1093/sleep/32.2.200

Plomin, R., & Von Stumm, S. (2022). Polygenic scores: Prediction versus explanation. Molecular Psychiatry, 27(1), 49–52. 10.1038/s41380-021-01348-y

Polderman, T. J. C., Benyamin, B., De Leeuw, C. A., Sullivan, P. F., Van Bochoven, A., Visscher, P. M., & Posthuma, D. (2015). Meta-analysis of the heritability of human traits based on fifty years of twin studies. Nature Genetics, 47(7), 702–709. 10.1038/ng.3285

Prather, A. A., Vogelzangs, N., & Penninx, B. W. J. H. (2015). Sleep duration, insomnia, and markers of systemic inflammation: Results from the Netherlands Study of Depression and Anxiety (NESDA). Journal of Psychiatric Research, 60, 95–102. 10.1016/j.jpsychires.2014.09.018

Sattar, N., & Preiss, D. (2017). Reverse Causality in Cardiovascular Epidemiological Research: More Common Than Imagined? Circulation, 135(24), 2369–2372. 10.1161/CIRCULATIONAHA.117.028307

Slavish, D. C., Asbee, J., Veeramachaneni, K., Messman, B. A., Scott, B., Sin, N. L., Taylor, D. J., & Dietch, J. R. (2021). The Cycle of Daily Stress and Sleep: Sleep Measurement Matters. Annals of Behavioral Medicine, 55(5), 413–423. 10.1093/abm/kaaa053

Stekhoven, D. J., & Bühlmann, P. (2012). MissForest—Non-parametric missing value imputation for mixed-type data. Bioinformatics, 28(1), 112–118. 10.1093/bioinformatics/btr597

Steptoe, A., Breeze, E., Banks, J., & Nazroo, J. (2013). Cohort Profile: The English Longitudinal Study of Ageing. International Journal of Epidemiology, 42(6), 1640–1648. 10.1093/ije/dys168

Steptoe, A., Hamer, M., & Chida, Y. (2007). The effects of acute psychological stress on circulating inflammatory factors in humans: A review and meta-analysis. *Brain*, Behavior, and Immunity, 21(7), 901–912. 10.1016/j.bbi.2007.03.011

Strain, T., Wijndaele, K., Sharp, S. J., Dempsey, P. C., Wareham, N., & Brage, S. (2020). Impact of follow-up time and analytical approaches to account for reverse causality on the association between physical activity and health outcomes in UK Biobank. International Journal of Epidemiology, 49(1), 162–172. 10.1093/ije/dyz212

Taillard, J., Gronfier, C., Bioulac, S., Philip, P., & Sagaspe, P. (2021). Sleep in Normal Aging, Homeostatic and Circadian Regulation and Vulnerability to Sleep Deprivation. Brain Sciences, 11(8), 1003. 10.3390/brainsci11081003

Tan, X., Shiyko, M. P., Li, R., Li, Y., & Dierker, L. (2012). A time-varying effect model for intensive longitudinal data. Psychological Methods, 17(1), 61–77. 10.1037/a0025814

Tomasik, J., Schiweck, C., & Drexhage, H. A. (2023). A Sticky Situation: The Link Between Peripheral Inflammation, Neuroinflammation, and Severe Mental Illness. Biological Psychiatry, 93(2), 107–109. 10.1016/j.biopsych.2022.10.017

Uy, J. P., Dieffenbach, M., Leschak, C. J., Eisenberger, N. I., Fuligni, A. J., & Galván, A. (2022). Sleep duration moderates the associations between immune markers and corticolimbic function during stress in adolescents. Neuropsychologia, 176, 108374. 10.1016/j.neuropsychologia.2022.108374

Vandekerckhove, M., Weiss, R., Schotte, C., Exadaktylos, V., Haex, B., Verbraecken, J., & Cluydts, R. (2011). The role of presleep negative emotion in sleep physiology. Psychophysiology, 48(12), 1738–1744. 10.1111/j.1469-8986.2011.01281.x

Vansteelandt, S., Dukes, O., & Martinussen, T. (2018). Survivor bias in Mendelian randomization analysis. Biostatistics, 19(4), 426–443. 10.1093/biostatistics/kxx050

Wardle, J., Chida, Y., Gibson, E. L., Whitaker, K. L., & Steptoe, A. (2011). Stress and Adiposity: A Meta-Analysis of Longitudinal Studies. Obesity, 19(4), 771–778. 10.1038/oby.2010.241

Whiteford, H. A., Degenhardt, L., Rehm, J., Baxter, A. J., Ferrari, A. J., Erskine, H. E., Charlson, F. J., Norman, R. E., Flaxman, A. D., Johns, N., Burstein, R., Murray, C. J., & Vos, T. (2013). Global burden of disease attributable to mental and substance use disorders: Findings from the Global Burden of Disease Study 2010. The Lancet, 382(9904), 1575– 1586. 10.1016/S0140-6736(13)61611-6

Wray, N. R., Lin, T., Austin, J., McGrath, J. J., Hickie, I. B., Murray, G. K., & Visscher, P. M. (2021). From Basic Science to Clinical Application of Polygenic Risk Scores: A Primer. JAMA Psychiatry, 78(1), 101. 10.1001/jamapsychiatry.2020.3049

Yap, Y., Slavish, D. C., Taylor, D. J., Bei, B., & Wiley, J. F. (2020). Bi-directional relations between stress and self-reported and actigraphy-assessed sleep: A daily intensive longitudinal study. Sleep, 43(3), zsz250. 10.1093/sleep/zsz250

Yuan, N., Chen, Y., Xia, Y., Dai, J., & Liu, C. (2019). Inflammation-related biomarkers in major psychiatric disorders: A cross-disorder assessment of reproducibility and specificity in 43 meta-analyses. Translational Psychiatry, 9(1), 233. 10.1038/s41398-019-0570-y

Zagaria, A., Ottaviani, C., Lombardo, C., & Ballesio, A. (2023). Perseverative Cognition as a Mediator Between Perceived Stress and Sleep Disturbance: A Structural Equation Modeling Meta-analysis (meta-SEM). Annals of Behavioral Medicine, 57(6), 463–471. 10.1093/abm/kaac064

