## Supplementary material for "Stress and Sleep Duration in Immune and Neuroendocrine Patterning. An Analytical Triangulation in ELSA": Full Supplement

***Figure S1.*** **Flow chart of missingness and the analytic sample for imputed data**

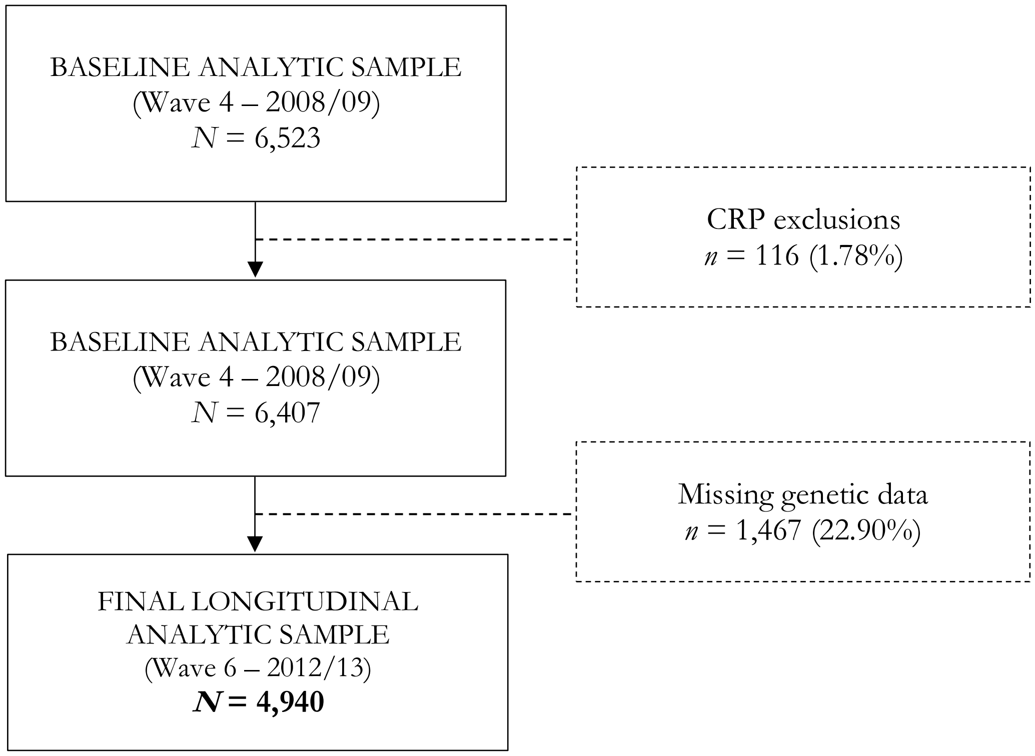

*Figure S2.* **Directed acyclic graph (DAG) conceptually representing causal effects between study variables**

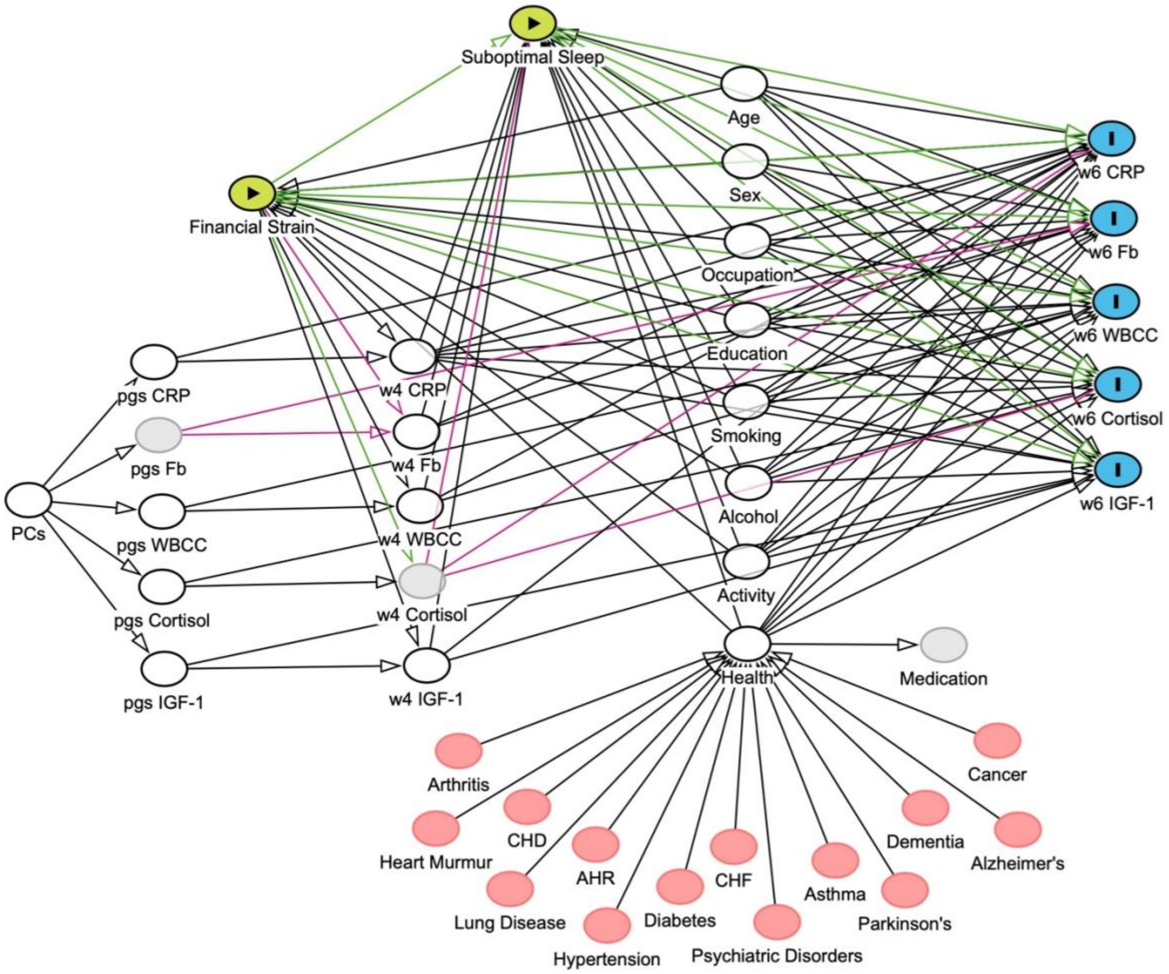


**KEY**

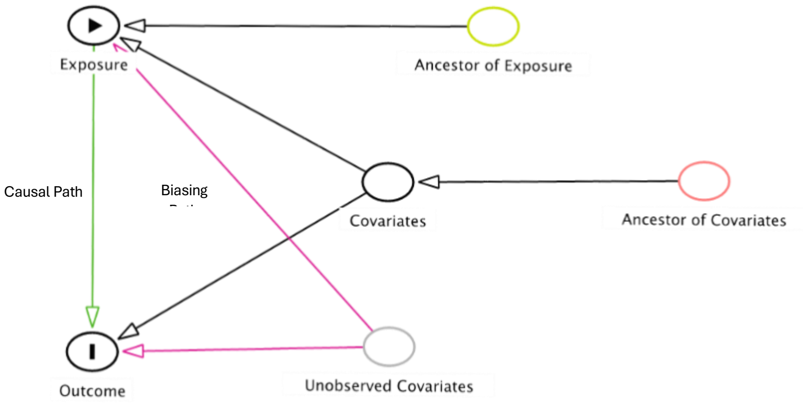

*Figure S3.* **The percentage of participants belonging to each immune and neuroendocrine biomarker profile with 95% confidence intervals for the wave 4 three-profile solution (N = 4,940)**

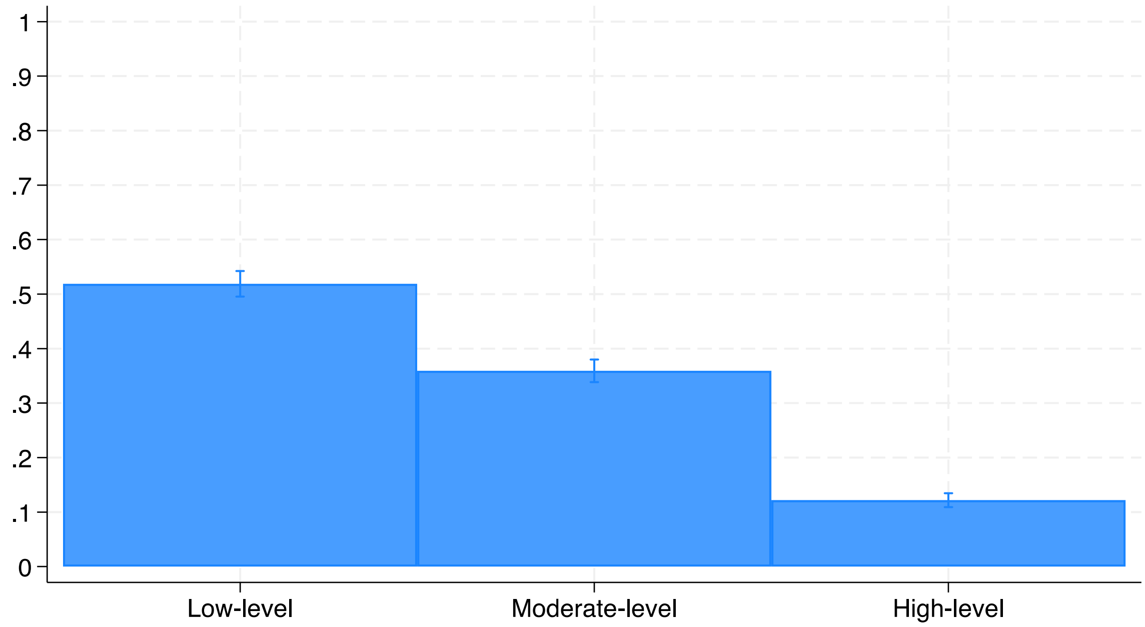


| **Profile** | **N** | **%** |
|---|---|---|
| **1** | 2,590 | 52.43 |
| **2** | 1,773 | 35.89 |
| **3** | 577 | 11.68 |

*Figure S4* [a-g]. **Predicted mean of immune and neuroendocrine biomarker levels for a one to seven profile solution for wave 6 (N = 4,940)**

a

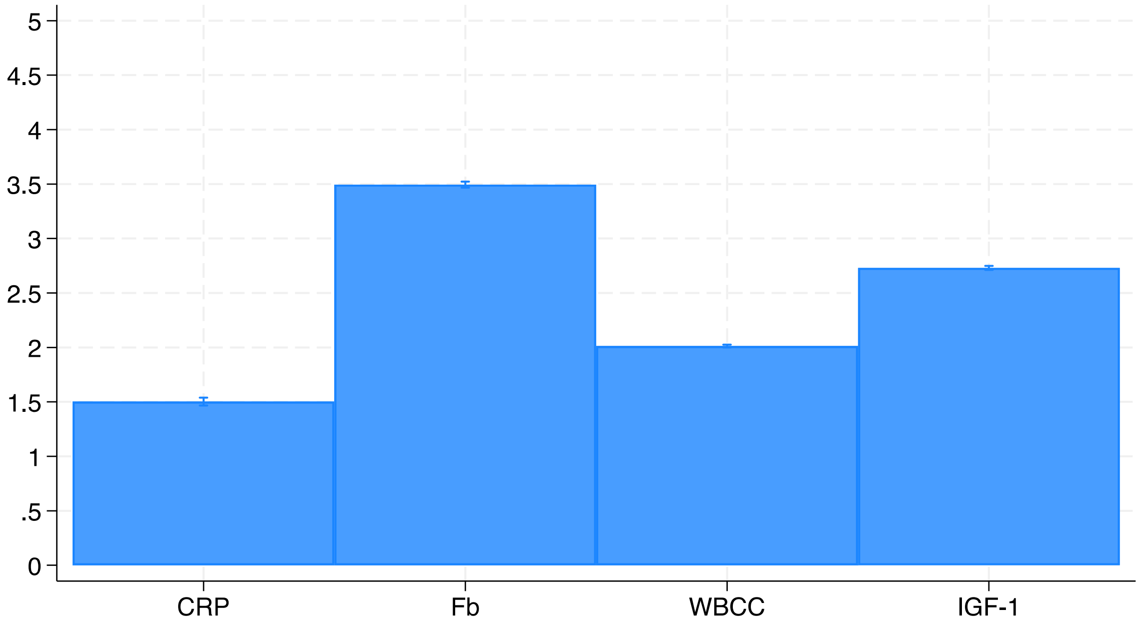


b

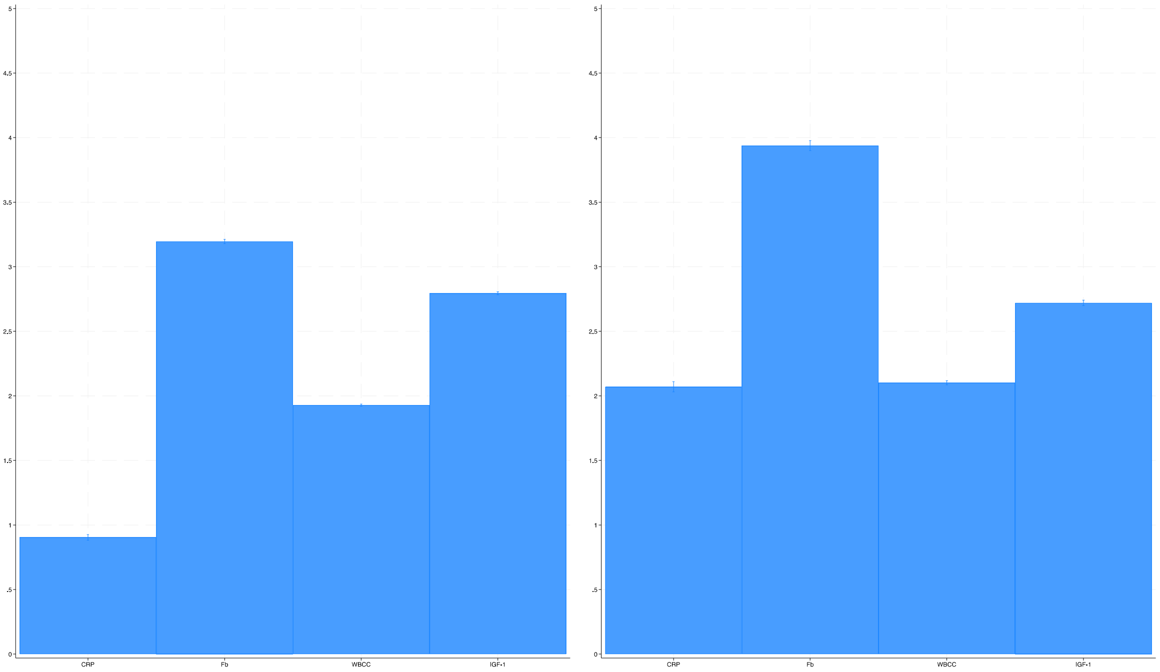

c

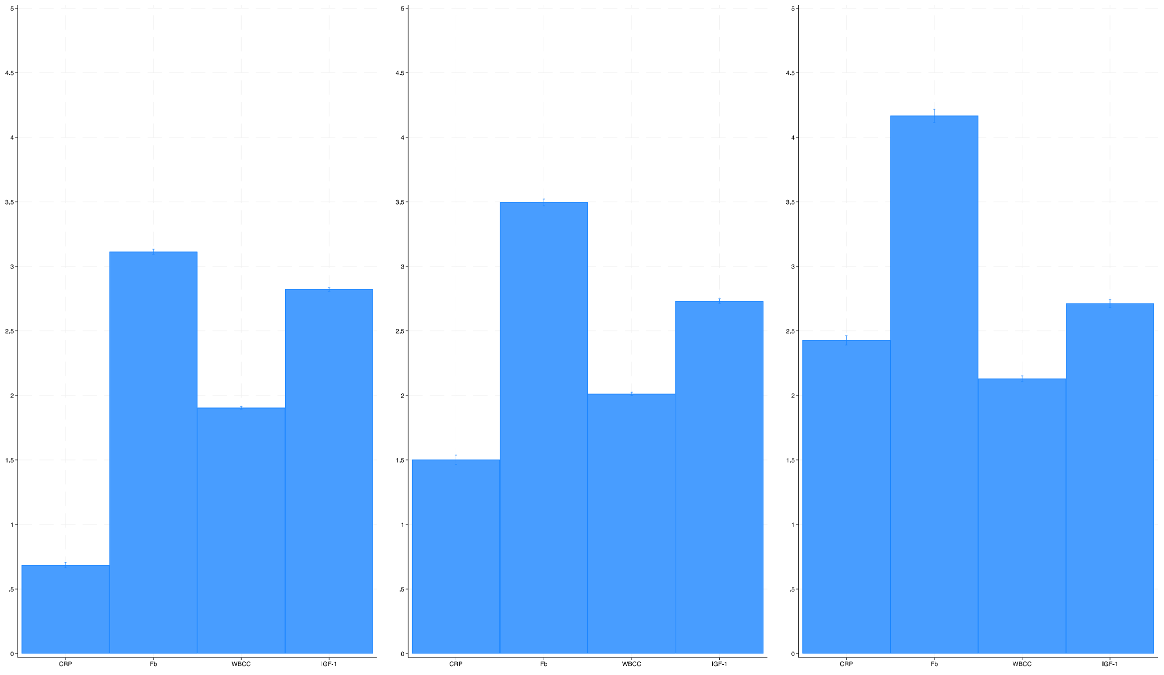
5
4.5
4
3.5
3
2.5
2
1.5
1
.5
0
CRP
Fb
WBCC
IGF-1

d

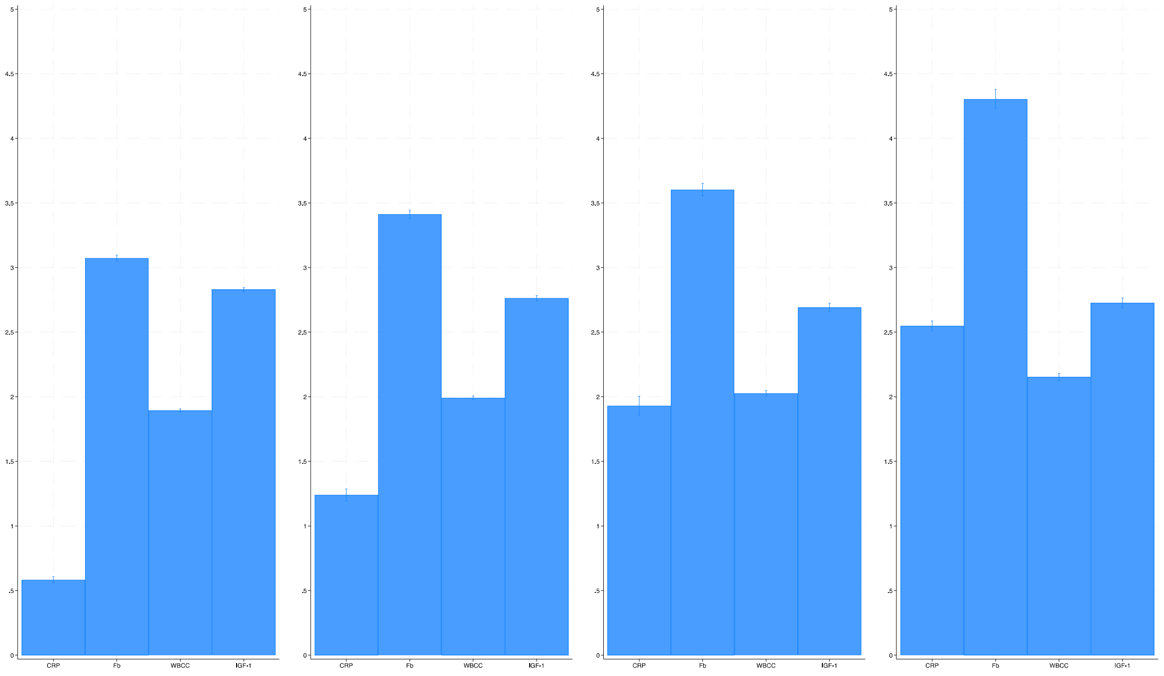
5
4.5
4
3.5
3
2.5
2
1.5
1
.5
0
CRP
Fb
WBCC
IGF-1

e

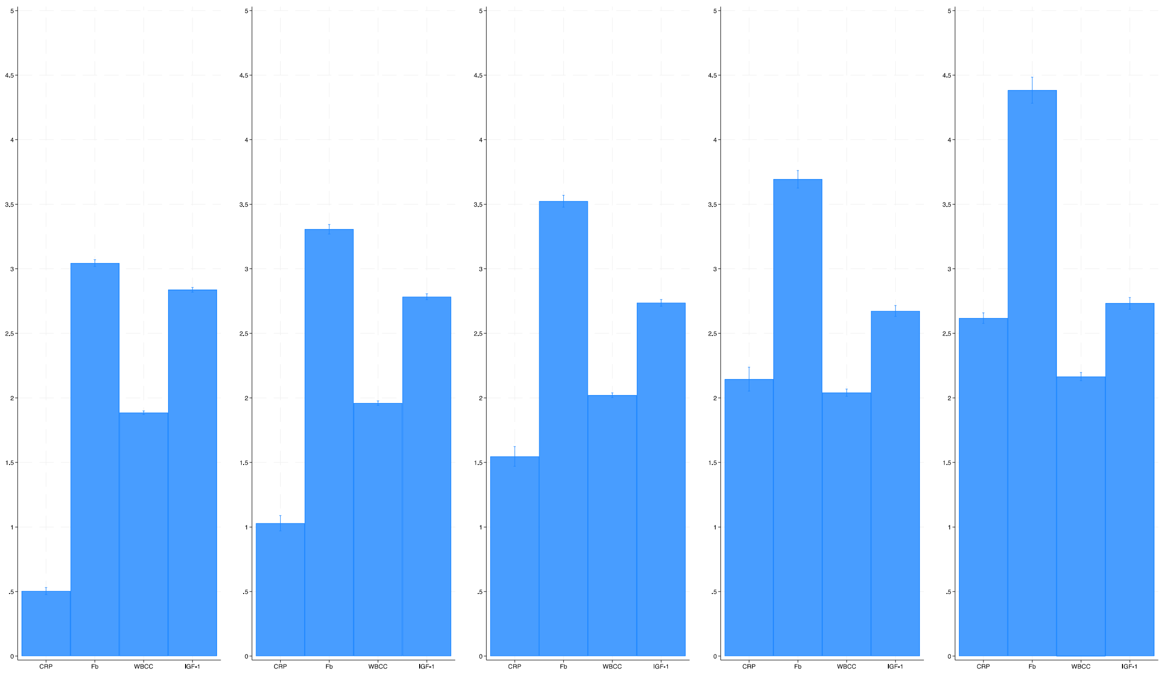
0
.5
1
1,5
2
2,5
3
3,5
4
4,5
5
CRP
Fb
WBCC
IGF-1

f

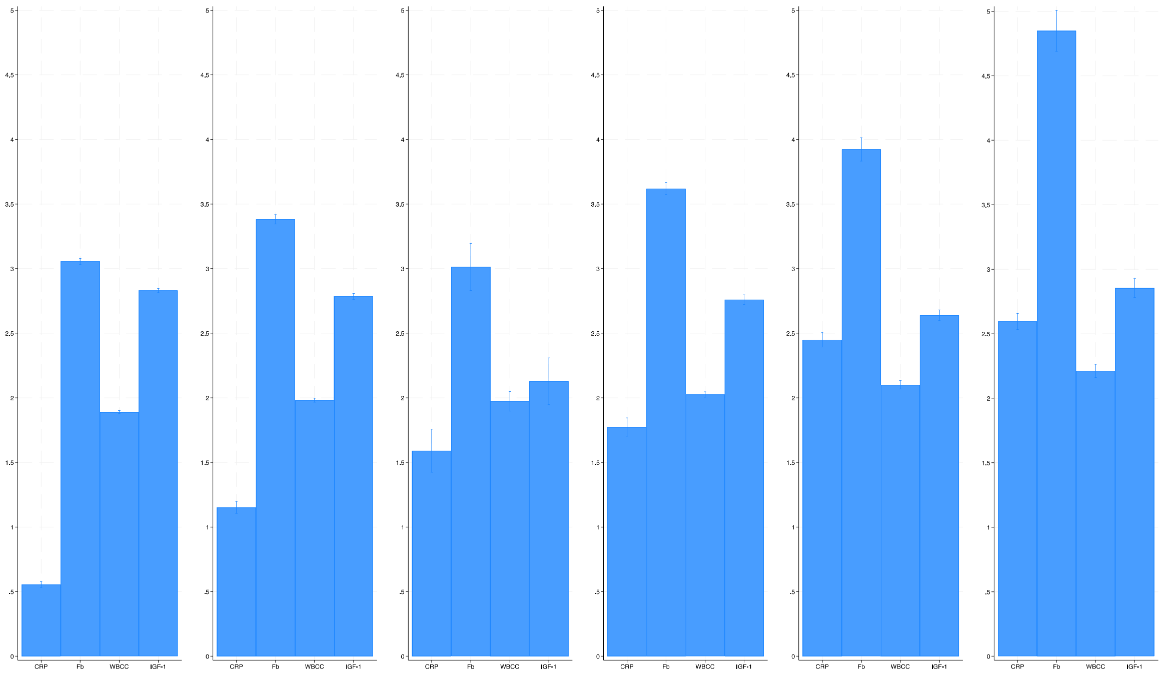
0
.5
1
1,5
2
2,5
3
3,5
4
4,5
5
CRP
Fb
WBCC
IGF-1

g

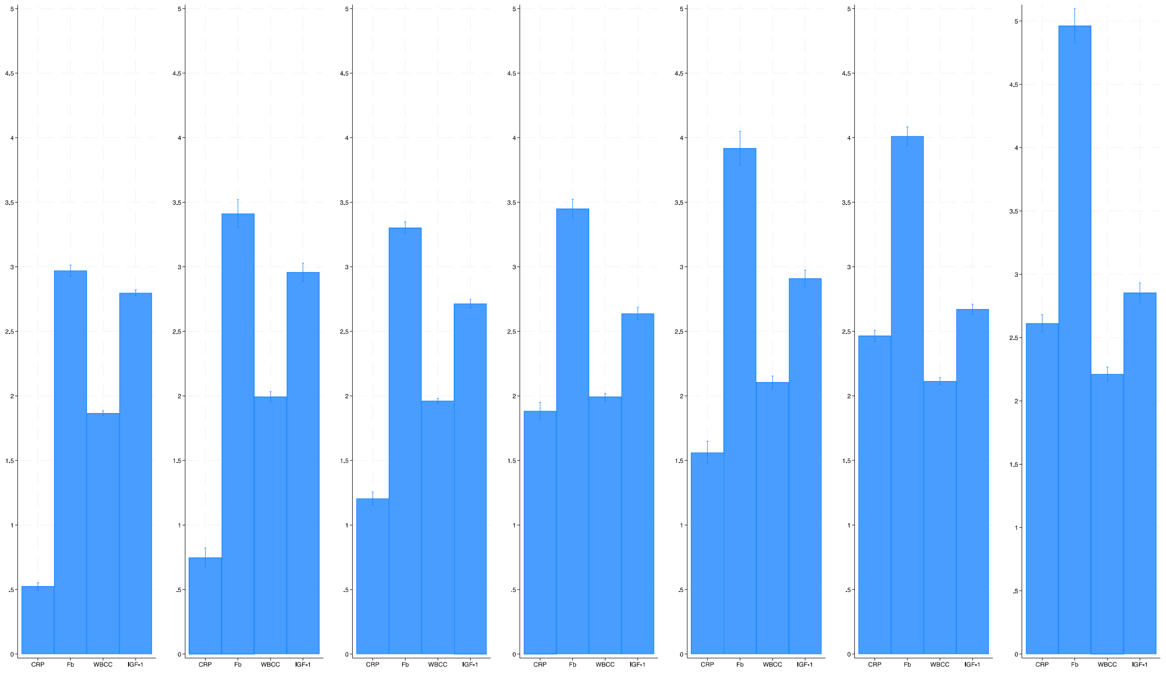

***Figure S5.*** **Akaike Information Criterion (AIC) and Bayesian Information Criterion (BIC) Values of Immune and Neuroendocrine Profiles to Assess Model Fit for wave 6**

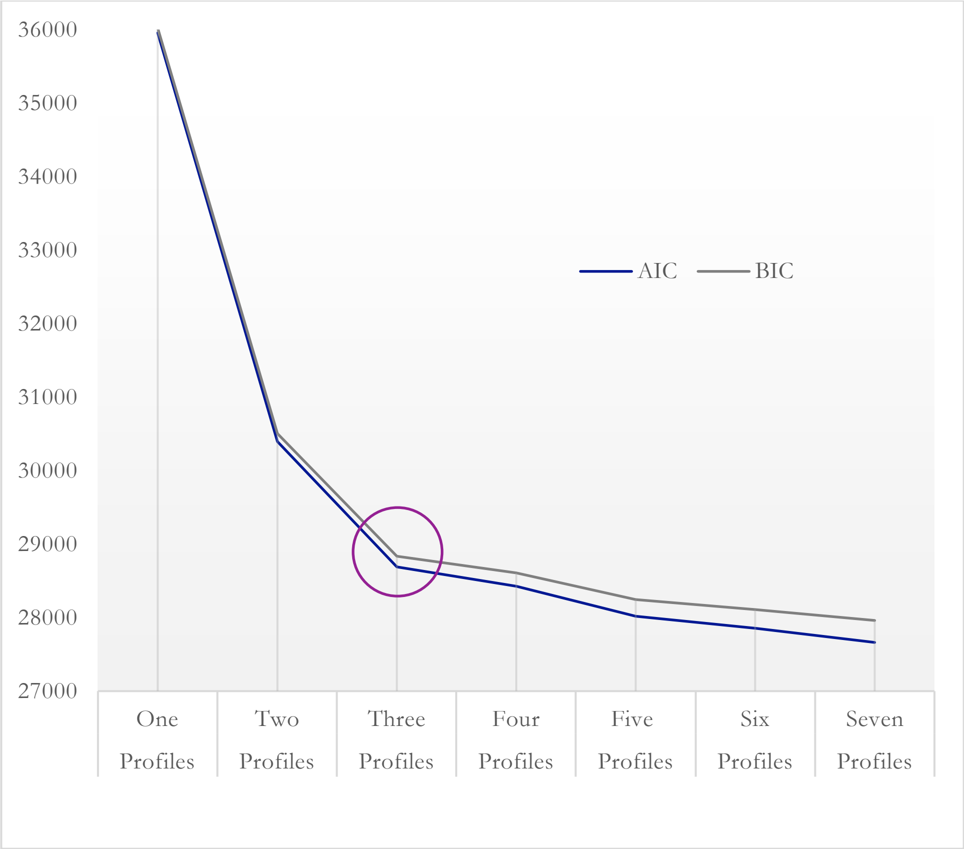

***Figure S6.*** **Entropy and Normalised Entropy Values of Immune and Neuroendocrine Biomarker Profiles to Assess Profile Quality for wave 6**

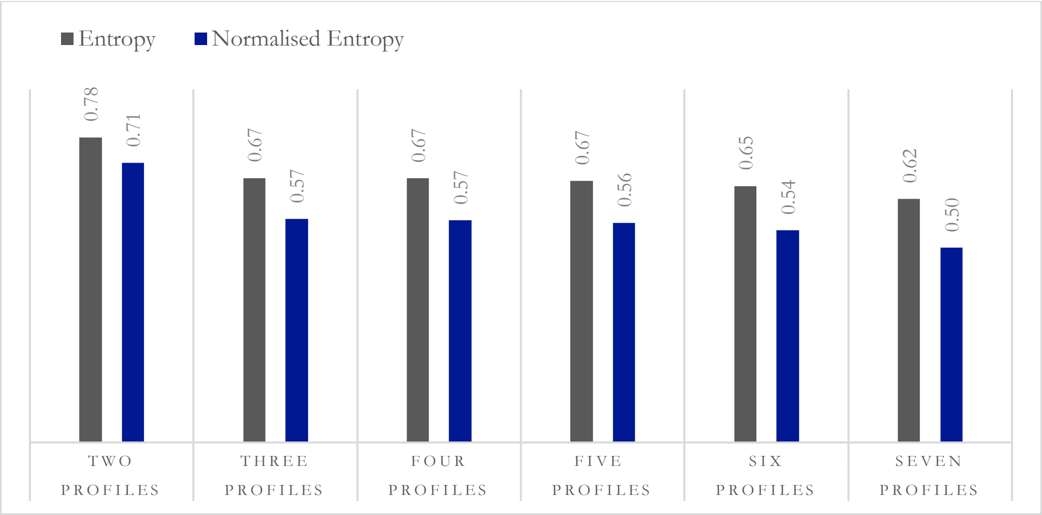

***Figure S7.*** **Mean Posterior Probabilities of Immune and Neuroendocrine Biomarker Profiles to Assess Membership Confidence (≥5%) for wave 6**

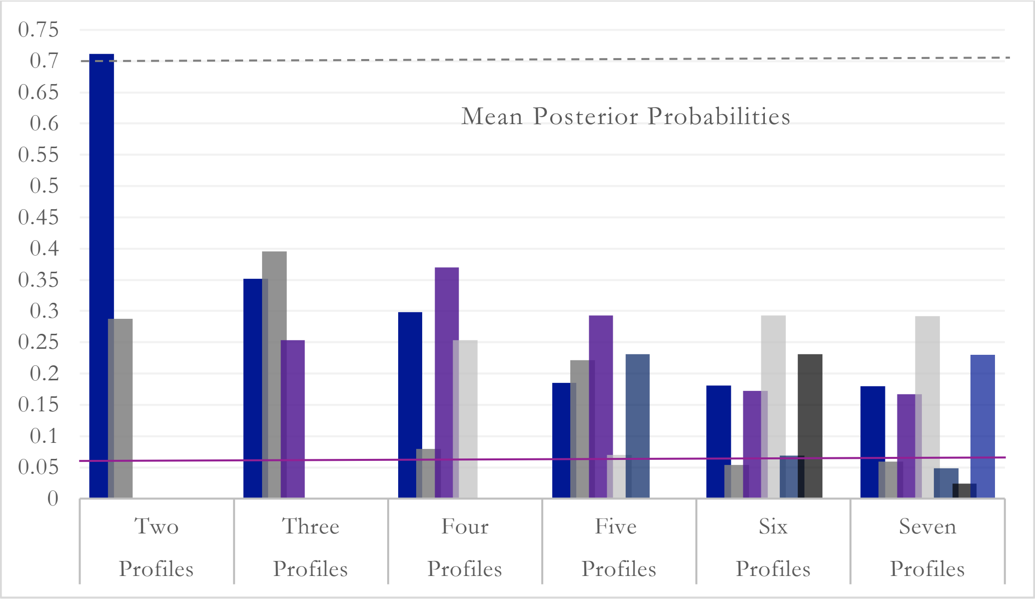

***Table S1.*** **The distribution of missing data, with a comparison between imputed, core, and complete case sample characteristics**

| Variable | | Missing Data | Imputed (N = 4,940) | | Core (N = 6,523) | | Complete Case (N = 1,305) | |
|---|---|---|---|---|---|---|---|---|
| | | N \| % | N / M (SD) | % / Range | N / M (SD) | % / Range | N / M (SD) | % / Range |
| Age | | 0 \| 0 | 66.3 (9.35) | 50-99 | 65.4 (9.43) | 50-99 | 64.53 (7.85) | 50-99 |
| Age (Binary) | < Md | 0 \| 0 | 2,436 | 49.31 | 3,205 | 49.13 | 628 | 48.12 |
| | ≥ Md | | 2,504 | 50.69 | 3,318 | 50.87 | 677 | 51.88 |
| Sex | Male | 0 \| 0 | 2,237 | 45.3 | 2,945 | 45.15 | 410 | 31.42 |
| | Female | | 2,703 | 54.7 | 3,578 | 54.85 | 895 | 68.58 |
| Education | Higher | 100 \| 0.93 | 1,589 | 32.2 | 2,116 | 32.44 | 445 | 34.10 |
| | Primary/Secondary/Tertiary | | 1,543 | 31.2 | 2,075 | 31.81 | 436 | 33.41 |
| | Alternative/No | | 1,808 | 36.6 | 2,332 | 35.75 | 424 | 32.49 |
| Wealth | Lowest | 1,160 \| 10.79 | 1,573 | 31.8 | 2,082 | 31.92 | 399 | 30.57 |
| | Middle | | 2,014 | 40.8 | 2,581 | 39.57 | 565 | 43.30 |
| | Highest | | 1,353 | 27.4 | 1,860 | 28.51 | 341 | 26.13 |
| Occupational Social Class | Managerial/Professional | 534 \| 4.97 | 1,793 | 36.3 | 2,434 | 37.31 | 475 | 36.40 |
| | Intermediate | | 1,264 | 25.6 | 1,641 | 25.16 | 352 | 26.97 |
| | Routine/Manual | | 1,883 | 38.1 | 2,448 | 37.53 | 478 | 36.63 |
| Smoking Status | Never/Ex-Smokers | 203 \| 1.89 | 4,312 | 87.3 | 5,673 | 86.97 | 1,146 | 87.82 |
| | Current Smoker | | 628 | 12.7 | 850 | 13.03 | 159 | 12.18 |
| Alcohol Consumption | <3 days a week | 1,597 \| 14.86 | 3,175 | 64.3 | 4,262 | 65.34 | 815 | 62.45 |
| | ≥3 days a week | | 1,765 | 35.7 | 2,261 | 34.66 | 490 | 37.55 |
| Physical Activity | Sedentary | 179 \| 1.67 | 1,340 | 27.1 | 1,786 | 27.38 | 304 | 23.30 |
| | Active | | 3,600 | 72.9 | 4,737 | 72.62 | 1,001 | 76.70 |
| Mobility | Mobile | 141 \| 1.31 | 2,678 | 54.2 | 3,422 | 52.46 | 687 | 52.64 |
| | Not Mobile | | 2,262 | 45.8 | 3,101 | 47.54 | 618 | 47.36 |
| Limiting Longstanding Illness | None | 8 \| 0.07 | 3,386 | 68.5 | 4,535 | 69.52 | 923 | 70.73 |
| | Present | | 1,554 | 31.5 | 1,988 | 30.48 | 382 | 29 |
| Health | No health condition | 132 \| 1.23 | 3,319 | 67.2 | 4,640 | 71.13 | 866 | 66.36 |
| | At least one health condition | | 1,621 | 32.8 | 1,883 | 28.87 | 439 | 33.64 |
| Body Mass Index (BMI) | <25, Underweight/Normal | 2,474 \| 23.02 | 1,312 | 26.6 | 1,769 | 27.12 | 354 | 27.13 |
| | 25-30, Overweight: Pre-obese | | 2,213 | 44.8 | 2,917 | 44.72 | 564 | 43.22 |
| | 30 or over, Obese | | 1,415 | 28.6 | 1,837 | 28.16 | 387 | 29.66 |
| CRP* (mg/L; Baseline) | | 4,502 \| 41.88 | 0.28 (0.46) | -0.70-1.30 | 0.28 (0.46) | -0.70-1.30 | 0.24 (0.46) | -0.70-1.29 |
| CRP* (mg/L; Follow-up) | | 5,625 \| 52.33 | 0.40 (0.48) | -1.00-1.30 | 0.39 (0.47) | -1.00-1.30 | 0.19 (0.43) | -1.00-1.29 |
| Fb (g/L; Baseline) | | 4,535 \| 42.19 | 3.38 (0.56) | 1.30-5.90 | 3.37 (0.56) | 1.30-5.90 | 3.32 (0.53) | 1.70-5.30 |
| Fb (g/L; Follow-up) | | 5,620 \| 52.28 | 3.12 (0.52) | 1.50-5.80 | 3.10 (0.52) | 1.30-5.80 | 2.95 (0.49) | 1.60-4.70 |
| WBCC* (nmol/L; Baseline) | | 4,471 \| 41.59 | 0.79 (0.13) | -0.10-1.50 | 0.79 (0.13) | -0.10-1.70 | 0.78 (0.13) | 0.15-1.50 |
| WBCC* (nmol/L; Follow-up) | | 5,571 \| 51.83 | 0.81 (0.11) | 0.34-1.51 | 0.81 (0.11) | 0.31-1.51 | 0.79 (0.13) | 0.39-1.51 |
| IGF-1* (nmol/L; Baseline) | | 4,441 \| 41.32 | 1.18 (0.16) | 0.30-1.81 | 1.17 (0.16) | 0.30-1.81 | 1.18 (0.15) | 0.60-1.66 |
| IGF-1* (nmol/L; Follow-up) | | | 1.18 (0.13) | 0.60-1.76 | 1.18 (0.13) | 0.60-1.76 | 1.19 (0.14) | 0.70-1.76 |
| Cortisol* (nmol/L; Follow-up) | | 6,556 \| 60.99 | 1.23 (0.65) | -0.85-2.82 | 0.39 (0.47) | -1.00-1.30 | 0.89 (0.59) | -0.85-2.82 |
| Stress (indexed by Financial Strain) | No Strain (0-60%) | 676 \| 6.29 | 4,099 | 83.0 | 5,407 | 82.89 | 1,049 | 80.38 |
| | Strain (61-100%) | | 841 | 17.0 | 1,116 | 17.11 | 256 | 19.62 |
| Sleep Duration | Short Sleep | 0 \| 0 | 627 | 12.7 | 828 | 12.69 | 181 | 13.72 |
| | Optimal Sleep | | 4,227 | 85.6 | 5,575 | 85.47 | 1,126 | 85.37 |
| | Long Sleep | | 86 | 1.70 | 120 | 1.84 | 12 | 0.91 |

Notes: ELSA, waves 4-6 (2008/09-2012/13); N = observations; M = Mean; Md = Median; % = percentage frequencies; SD = standard deviations; < = less than; ≥ = greater than or equal to; OSC = occupational social class; CRP = C-reactive protein; Fb = Fibrinogen; WBCC = White Blood Cell Counts; IGF-1 = Insulin-growth factor-1; Cortisol = Hair Cortisol; * Log-transformed variable; I-N = Immune and Neuroendocrine.

***Table S2.*** **Correlations between immune and neuroendocrine biomarkers**

| | CRP | Fb | WBCC | Cortisol | IGF-1 |
|---|---|---|---|---|---|
| CRP | 1 | | | | |
| Fb | **0.7065*** | 1 | | | |
| | <0.001 | | | | |
| WBCC | **0.4477*** | **0.4122*** | 1 | | |
| | <0.001 | <0.001 | | | |
| Cortisol | **0.2811*** | **0.1893*** | **0.2317*** | 1 | |
| | <0.001 | <0.001 | <0.001 | | |
| IGF-1 | **-0.1671*** | -0.011 | -0.006 | 0.004 | 1 |
| | <0.001 | 0.356 | 0.627 | 0.755 | |

Notes: C-reactive protein (CRP); fibrinogen (Fb); white blood cell counts (WBCC); insulin growth factor-1 (IGF-1) hair cortisol (Cortisol); * Significant at the 0.001 level

***Table S3.*** **Seven Profile LPA model fit indices and predicted probability of profile membership**

| **Criteria** | **One Profile** | **Two Profiles** | **Three Profiles** | **Four Profiles** | **Five Profiles** | **Six Profiles** | **Seven Profiles** |
|---|---|---|---|---|---|---|---|
| AIC | 35960.37 | 30398.85 | 28688.66 | 28428.96 | 28023.89 | 27852.44 | 27662.91 |
| AIC Difference (N) | - | 5561.52 | 1710.19 | 259.70 | 405.07 | 171.45 | 189.53 |
| AIC Difference (%) | - | 18.30 | 5.96 | 0.91 | 1.45 | 0.62 | 0.69 |
| BIC | 36025.42 | 30502.93 | 28831.77 | 28611.10 | 28245.06 | 28112.65 | 27962.14 |
| BIC Difference (N) | - | 5522.49 | 1671.16 | 220.67 | 366.04 | 132.41 | 150.51 |
| BIC Difference (%) | - | 18.10 | 5.80 | 0.77 | 1.30 | 0.47 | 0.54 |
| aBIC | 35993.64 | 30452.09 | 28761.86 | 28522.13 | 28137.02 | 27985.54 | 27815.97 |
| aBIC Difference (N) | - | 5541.55 | 1690.23 | 239.74 | 385.11 | 151.48 | 169.57 |
| aBIC Difference (%) | - | 18.20 | 5.88 | 0.84 | 1.37 | 0.54 | 0.61 |
| Entropy | - | 0.91 | 0.83 | 0.79 | 0.83 | 0.86 | 0.86 |
| Normalised Entropy | - | 0.88 | 0.77 | 0.71 | 0.77 | 0.82 | 0.81 |
| M Posterior Probabilities (SE) | - | .712 (.007) | .352 (.008) | .298 (.013) | .185 (.008) | .181 (.008) | .180 (.008) |
| | | .288 (.007) | .395 (.008) | .079 (.014) | .221 (.008) | .054 (.012) | .059 (.011) |
| | | | .254 (.006) | .370 (.009) | .293 (.008) | .172 (.114) | .167 (.013) |
| | | | | .253 (.006) | .070 (.005) | .293 (.008) | .292 (.008) |
| | | | | | .231 (.006) | .069 (.005) | .048 (.006) |
| | | | | | | .231 (.006) | .024 (.004) |
| | | | | | | | .230 (.006) |
| N classes >5% | Yes | Yes | Yes | Yes | Yes | No | No |

Notes: AIC = Akaike information criterion; BIC = Bayesian information criterion; aBIC = adjusted Bayesian information criterion; N = number of observations; M = mean; SE = standard errors.

***Table S4.*** **Longitudinal associations of stress with immune and neuroendocrine biomarker profiles stratified by median age (65 years)**

| **Adjustments** | **Binary Stress Score \| < Md Age (N=2,436)** | | | | | **Binary Stress Score \| ≥ Md Age (N=2,504)** | | | | |
|---|---|---|---|---|---|---|---|---|---|---|
| | **RRR** | **SE** | **95% CI** | | ***p*** | **RRR** | **SE** | **95% CI** | | ***p*** |
| ***Moderate-risk* Profile** | | | | | | | | | | |
| Model 1: *Unadjusted* | 1.39 | 0.17 | 1.09 | 1.78 | 0.008 | 1.12 | 0.15 | 0.86 | 1.46 | 0.383 |
| Model 2: *Model 1 + baseline biomarkers* [a] | 1.39 | 0.18 | 1.08 | 1.80 | 0.011 | 1.10 | 0.15 | 0.84 | 1.43 | 0.498 |
| Model 3: *Model 2 + demographics & genetics* [b] | 1.38 | 0.18 | 1.07 | 1.79 | 0.014 | 1.19 | 0.17 | 0.90 | 1.56 | 0.221 |
| Model 4: *Fully Adjusted* [c] | 1.30 | 0.18 | 1.00 | 1.70 | 0.050 | 1.08 | 0.15 | 0.81 | 1.42 | 0.614 |
| ***High-risk* Profile** | | | | | | | | | | |
| Model 1: *Unadjusted* | 1.97 | 0.28 | 1.50 | 2.60 | <0.001 | 1.31 | 0.18 | 0.99 | 1.72 | 0.056 |
| Model 2: *Model 1 + baseline biomarkers* [a] | 1.98 | 0.33 | 1.43 | 2.75 | <0.001 | 1.24 | 0.20 | 0.91 | 1.69 | 0.176 |
| Model 3: *Model 2 + demographics & genetics* [b] | 1.97 | 0.33 | 1.42 | 2.75 | <0.001 | 1.40 | 0.23 | 1.02 | 1.93 | 0.040 |
| Model 4: *Fully Adjusted* [c] | 1.70 | 0.30 | 1.20 | 2.41 | 0.003 | 1.23 | 0.21 | 0.88 | 1.71 | 0.232 |

Notes: The *low-risk* group is the reference; < = less than; ≥ = greater than or equal to; Md = Median; RRR = relative risk ratio; SE = standard errors; CI = confidence interval; *p* = significance value.

a Baseline biomarkers: C-reactive protein (CRP); fibrinogen; insulin-growth factor-1 (IGF-1).

b Demographic and genetic variables: age; sex; 10 principal components (PCs); CRP polygenic score (PGS); cortisol PGS; IGF-1 PGS.

c All variables: CRP; fibrinogen; IGF-1; age; sex; 10 PCs; CRP PGS; cortisol PGS; IGF-1 PGS; education; occupational social status; smoking status; alcohol consumption; physical activity; health (i.e., chronic lung disease; coronary heart disease; abnormal heart rhythm; heart murmur; congestive heart failure; angina; hypertension; diabetes; cancer; Parkinson's; Alzheimer's; dementia; asthma; arthritis; osteoporosis; psychiatric disorder).

***Table S5.*** **Longitudinal associations of suboptimal sleep with immune and neuroendocrine biomarker profiles stratified by median age (65 years)**

| **Adjustments** | **Short Sleep \| < Md Age (N=2,436)** | | | | | **Short Sleep \| ≥ Md Age (N=2,504)** | | | | |
|---|---|---|---|---|---|---|---|---|---|---|
| | **RRR** | **SE** | **95% CI** | | ***p*** | **RRR** | **SE** | **95% CI** | | ***p*** |
| ***Moderate-risk*** **Profile** | | | | | | | | | | |
| Model 1: *Unadjusted* | 1.09 | 0.16 | 0.82 | 1.45 | 0.547 | 1.17 | 0.17 | 0.88 | 1.56 | 0.281 |
| Model 2: *Model 1 + baseline biomarkers* [a] | 0.95 | 0.15 | 0.70 | 1.29 | 0.745 | 1.15 | 0.17 | 0.86 | 1.54 | 0.335 |
| Model 3: *Model 2 + demographics & genetics* [b] | 0.93 | 0.14 | 0.69 | 1.26 | 0.636 | 1.07 | 0.16 | 0.79 | 1.44 | 0.664 |
| Model 4: *Fully Adjusted* [c] | 0.87 | 0.14 | 0.64 | 1.19 | 0.387 | 0.93 | 0.15 | 0.68 | 1.26 | 0.629 |
| ***High-risk*** **Profile** | | | | | | | | | | |
| Model 1: *Unadjusted* | 1.61 | 0.26 | 1.18 | 2.21 | 0.003 | 1.29 | 0.20 | 0.96 | 1.75 | 0.093 |
| Model 2: *Model 1 + baseline biomarkers* [a] | 1.15 | 0.23 | 0.79 | 1.69 | 0.471 | 1.29 | 0.22 | 0.92 | 1.81 | 0.148 |
| Model 3: *Model 2 + demographics & genetics* [b] | 1.14 | 0.23 | 0.77 | 1.69 | 0.504 | 1.16 | 0.21 | 0.82 | 1.65 | 0.410 |
| Model 4: *Fully Adjusted* [c] | 0.89 | 0.19 | 0.59 | 1.35 | 0.594 | 0.84 | 0.16 | 0.58 | 1.21 | 0.351 |
| | **Long Sleep \| < Md Age (N=2,436)** | | | | | **Long Sleep \| ≥ Md Age (N=2,504)** | | | | |
| ***Moderate-risk*** **Profile** | | | | | | | | | | |
| Model 1: *Unadjusted* | 1.98 | 1.01 | 0.73 | 5.38 | 0.180 | 1.22 | 0.47 | 0.57 | 2.60 | 0.604 |
| Model 2: *Model 1 + baseline biomarkers* [a] | 1.54 | 0.83 | 0.53 | 4.43 | 0.426 | 1.20 | 0.47 | 0.56 | 2.57 | 0.648 |
| Model 3: *Model 2 + demographics & genetics* [b] | 1.53 | 0.83 | 0.53 | 4.43 | 0.437 | 0.94 | 0.38 | 0.43 | 2.06 | 0.876 |
| Model 4: *Fully Adjusted* [c] | 1.39 | 0.77 | 0.47 | 4.11 | 0.554 | 0.86 | 0.35 | 0.38 | 1.92 | 0.706 |
| ***High-risk*** **Profile** | | | | | | | | | | |
| Model 1: *Unadjusted* | 3.70 | 1.92 | 1.34 | 10.25 | 0.012 | 2.80 | 1.00 | 1.39 | 5.63 | 0.004 |
| Model 2: *Model 1 + baseline biomarkers* [a] | 1.79 | 1.14 | 0.52 | 6.23 | 0.358 | 2.68 | 1.07 | 1.22 | 5.88 | 0.014 |
| Model 3: *Model 2 + demographics & genetics* [b] | 2.00 | 1.28 | 0.58 | 6.99 | 0.275 | 1.78 | 0.75 | 0.78 | 4.07 | 0.173 |
| Model 4: *Fully Adjusted* [c] | 1.44 | 0.95 | 0.40 | 5.22 | 0.575 | 1.35 | 0.59 | 0.57 | 3.20 | 0.502 |

Notes: The *low-risk* group is the reference; < = less than; ≥ = greater than or equal to; Md = Median; RRR = relative risk ratio; SE = standard errors; CI = confidence interval; *p* = significance value.

a Baseline biomarkers: C-reactive protein (CRP); fibrinogen; insulin-growth factor-1 (IGF-1).

b Demographic and genetic variables: age; sex; 10 principal components (PCs); CRP polygenic score (PGS); cortisol PGS; IGF-1 PGS.

c All variables: CRP; fibrinogen; IGF-1; age; sex; 10 PCs; CRP PGS; cortisol PGS; IGF-1 PGS; education; occupational social status; smoking status; alcohol consumption; physical activity; health (i.e., chronic lung disease; coronary heart disease; abnormal heart rhythm; heart murmur; congestive heart failure; angina; hypertension; diabetes; cancer; Parkinson's; Alzheimer's; dementia; asthma; arthritis; osteoporosis; psychiatric disorder).

***Table S6.*** **Longitudinal associations of stress with immune and neuroendocrine biomarker profiles stratified by sex**

| Adjustments | Binary Stress Score \| Male (N=2,237) | | | | | Binary Stress Score \| Female (N=2,703) | | | | |
|---|---|---|---|---|---|---|---|---|---|---|
| | RRR | SE | 95% CI | | *p* | RRR | SE | 95% CI | | *p* |
| ***Moderate-risk*** **Profile** | | | | | | | | | | |
| Model 1: *Unadjusted* | 1.18 | 0.17 | 0.88 | 1.57 | 0.267 | 1.28 | 0.15 | 1.02 | 1.62 | 0.033 |
| Model 2: *Model 1 + baseline biomarkers* [a] | 1.20 | 0.18 | 0.90 | 1.60 | 0.225 | 1.23 | 0.15 | 0.97 | 1.56 | 0.093 |
| Model 3: *Model 2 + demographics & genetics* [b] | 1.25 | 0.19 | 0.93 | 1.67 | 0.145 | 1.27 | 0.16 | 0.99 | 1.61 | 0.058 |
| Model 4: *Fully Adjusted* [c] | 1.18 | 0.18 | 0.87 | 1.60 | 0.289 | 1.15 | 0.15 | 0.90 | 1.48 | 0.275 |
| ***High-risk*** **Profile** | | | | | | | | | | |
| Model 1: *Unadjusted* | 1.78 | 0.27 | 1.32 | 2.40 | <0.001 | 1.42 | 0.18 | 1.10 | 1.83 | 0.006 |
| Model 2: *Model 1 + baseline biomarkers* [a] | 1.91 | 0.34 | 1.35 | 2.69 | <0.001 | 1.31 | 0.20 | 0.97 | 1.76 | 0.074 |
| Model 3: *Model 2 + demographics & genetics* [b] | 2.02 | 0.36 | 1.42 | 2.87 | <0.001 | 1.41 | 0.22 | 1.04 | 1.91 | 0.027 |
| Model 4: *Fully Adjusted* [c] | 1.68 | 0.32 | 1.16 | 2.43 | 0.006 | 1.23 | 0.20 | 0.90 | 1.68 | 0.203 |

Notes: The *low-risk* group is the reference; < = less than; ≥ = greater than or equal to; RRR = relative risk ratio; SE = standard errors; CI = confidence interval; *p* = significance value.
a Baseline biomarkers: C-reactive protein (CRP); fibrinogen; insulin-growth factor-1 (IGF-1).
b Demographic and genetic variables: age; sex; 10 principal components (PCs); CRP polygenic score (PGS); cortisol PGS; IGF-1 PGS.
c All variables: CRP; fibrinogen; IGF-1; age; sex; 10 PCs; CRP PGS; cortisol PGS; IGF-1 PGS; education; occupational social status; smoking status; alcohol consumption; physical activity; health (i.e., chronic lung disease; coronary heart disease; abnormal heart rhythm; heart murmur; congestive heart failure; angina; hypertension; diabetes; cancer; Parkinson's; Alzheimer's; dementia; asthma; arthritis; osteoporosis; psychiatric disorder).

***Table S7.*** **Longitudinal associations of suboptimal sleep with immune and neuroendocrine biomarker profiles stratified by sex**

| Adjustments | Short Sleep \| Male (N=2,237) | | | | | Short Sleep \| Female (N=2,703) | | | | |
|---|---|---|---|---|---|---|---|---|---|---|
| | **RRR** | **SE** | **95% CI** | | ***p*** | **RRR** | **SE** | **95% CI** | | ***p*** |
| ***Moderate-risk* Profile** | | | | | | | | | | |
| Model 1: *Unadjusted* | 1.04 | 0.17 | 0.75 | 1.43 | 0.837 | 1.20 | 0.16 | 0.93 | 1.55 | 0.162 |
| Model 2: *Model 1 + baseline biomarkers* [a] | 1.00 | 0.17 | 0.72 | 1.40 | 0.981 | 1.12 | 0.15 | 0.85 | 1.46 | 0.427 |
| Model 3: *Model 2 + demographics & genetics* [b] | 0.95 | 0.16 | 0.68 | 1.33 | 0.766 | 1.06 | 0.15 | 0.81 | 1.38 | 0.694 |
| Model 4: *Fully Adjusted* [c] | 0.83 | 0.15 | 0.58 | 1.18 | 0.293 | 0.94 | 0.13 | 0.71 | 1.24 | 0.669 |
| ***High-risk* Profile** | | | | | | | | | | |
| Model 1: *Unadjusted* | 1.41 | 0.25 | 1.00 | 1.99 | 0.050 | 1.46 | 0.21 | 1.10 | 1.92 | 0.008 |
| Model 2: *Model 1 + baseline biomarkers* [a] | 1.33 | 0.27 | 0.89 | 1.97 | 0.166 | 1.23 | 0.21 | 0.89 | 1.71 | 0.212 |
| Model 3: *Model 2 + demographics & genetics* [b] | 1.21 | 0.25 | 0.80 | 1.82 | 0.365 | 1.17 | 0.20 | 0.84 | 1.63 | 0.362 |
| Model 4: *Fully Adjusted* [c] | 0.77 | 0.17 | 0.50 | 1.19 | 0.233 | 0.93 | 0.17 | 0.66 | 1.32 | 0.697 |
| | **Long Sleep \| Male (N=2,237)** | | | | | **Long Sleep \| Female (N=2,703)** | | | | |
| ***Moderate-risk* Profile** | | | | | | | | | | |
| Model 1: *Unadjusted* | 1.46 | 0.77 | 0.52 | 4.12 | 0.476 | 1.56 | 0.59 | 0.75 | 3.28 | 0.237 |
| Model 2: *Model 1 + baseline biomarkers* [a] | 1.41 | 0.76 | 0.49 | 4.03 | 0.522 | 1.38 | 0.54 | 0.64 | 2.99 | 0.415 |
| Model 3: *Model 2 + demographics & genetics* [b] | 1.13 | 0.61 | 0.39 | 3.27 | 0.821 | 1.11 | 0.45 | 0.51 | 2.44 | 0.796 |
| Model 4: *Fully Adjusted* [c] | 1.02 | 0.56 | 0.35 | 3.01 | 0.969 | 0.99 | 0.41 | 0.44 | 2.21 | 0.982 |
| ***High-risk* Profile** | | | | | | | | | | |
| Model 1: *Unadjusted* | 4.19 | 2.02 | 1.63 | 10.78 | 0.003 | 3.12 | 1.15 | 1.52 | 6.43 | 0.002 |
| Model 2: *Model 1 + baseline biomarkers* [a] | 4.51 | 2.41 | 1.58 | 12.86 | 0.005 | 2.01 | 0.89 | 0.85 | 4.79 | 0.114 |
| Model 3: *Model 2 + demographics & genetics* [b] | 3.45 | 1.88 | 1.19 | 10.06 | 0.023 | 1.47 | 0.66 | 0.61 | 3.56 | 0.395 |
| Model 4: *Fully Adjusted* [c] | 2.49 | 1.43 | 0.81 | 7.65 | 0.111 | 1.11 | 0.51 | 0.45 | 2.75 | 0.820 |

Notes: The *low-risk* group is the reference; < = less than; ≥ = greater than or equal to; RRR = relative risk ratio; SE = standard errors; CI = confidence interval; *p* = significance value.

a Baseline biomarkers: C-reactive protein (CRP); fibrinogen; insulin-growth factor-1 (IGF-1).

b Demographic and genetic variables: age; sex; 10 principal components (PCs); CRP polygenic score (PGS); cortisol PGS; IGF-1 PGS.

c All variables: CRP; fibrinogen; IGF-1; age; sex; 10 PCs; CRP PGS; cortisol PGS; IGF-1 PGS; education; occupational social status; smoking status; alcohol consumption; physical activity; health (i.e., chronic lung disease; coronary heart disease; abnormal heart rhythm; heart murmur; congestive heart failure; angina; hypertension; diabetes; cancer; Parkinson's; Alzheimer's; dementia; asthma; arthritis; osteoporosis; psychiatric disorder).

***Table S8.*** **Longitudinal associations of stress with immune and neuroendocrine profiles, adjusted for BMI**

| **Adjustments** | **Stress** | | | | |
|---|---|---|---|---|---|
| | **RRR** | **SE** | **95% CI** | | ***p*** |
| ***Moderate-risk* Profile** | | | | | |
| Model 1: *Unadjusted* | 1.26 | 0.12 | 1.05 | 1.50 | 0.012 |
| Model 2: *Model 1 + baseline biomarkers* [a] | 1.23 | 0.12 | 1.03 | 1.48 | 0.025 |
| Model 3: *Model 2 + demographics & genetics* [b] | 1.28 | 0.12 | 1.060 | 1.54 | 0.010 |
| Model 4: *Fully Adjusted* [c] | 1.18 | 0.12 | 0.97 | 1.43 | 0.099 |
| ***High-risk* Profile** | | | | | |
| Model 1: *Unadjusted* | 1.57 | 0.15 | 1.30 | 1.90 | <0.001 |
| Model 2: *Model 1 + baseline biomarkers* [a] | 1.52 | 0.17 | 1.21 | 1.90 | <0.001 |
| Model 3: *Model 2 + demographics & genetics* [b] | 1.65 | 0.19 | 1.308 | 2.07 | <0.001 |
| Model 4: *Fully Adjusted* [c] | 1.42 | 0.17 | 1.12 | 1.80 | 0.004 |

Notes: The *low-risk* group is the reference; RRR = relative risk ratio; SE = standard errors; CI = confidence interval; *p* = significance value.

a Baseline immune and neuroendocrine profiles.

b Demographic and genetic variables: age; sex; 10 principal components (PCs); C-reactive Protein (CRP) polygenic score (PGS); White Blood Cell Counts (WBCC) PGS; Insulin Growth Factor-1 (IGF-1) PGS; [Hair] Cortisol PGS; Sleep Duration PGS.

c All variables: Baseline immune and neuroendocrine profiles; age; sex; 10 PCs; CRP PGS; WBCC PGS; IGF-1 PGS; Cortisol PGS; education; wealth; occupational social status; smoking status; alcohol consumption; physical activity; mobility; limiting longstanding illness; health (i.e., chronic lung disease; coronary heart disease; abnormal heart rhythm; heart murmur; congestive heart failure; angina; hypertension; diabetes; cancer; Parkinson's; Alzheimer's; dementia; asthma; arthritis; osteoporosis; psychiatric disorder); BMI.

***Table S9.*** **Longitudinal associations of suboptimal sleep with immune and neuroendocrine profiles, adjusted for BMI**

| **Adjustments** | **Short Sleep** | | | | |
|---|---|---|---|---|---|
| | **RRR** | **SE** | **95% CI** | | ***p*** |
| ***Moderate-risk* Profile** | | | | | |
| Model 1: *Unadjusted* | 1.15 | 0.12 | 0.94 | 1.41 | 0.171 |
| Model 2: *Model 1 + baseline biomarkers* [a] | 1.08 | 0.11 | 0.88 | 1.33 | 0.447 |
| Model 3: *Model 2 + demographics & genetics* [b] | 1.01 | 0.11 | 0.82 | 1.25 | 0.907 |
| Model 4: *Fully Adjusted* [c] | 0.89 | 0.10 | 0.71 | 1.10 | 0.281 |
| ***High-risk* Profile** | | | | | |
| Model 1: *Unadjusted* | 1.45 | 0.16 | 1.17 | 1.80 | 0.001 |
| Model 2: *Model 1 + baseline biomarkers* [a] | 1.26 | 0.16 | 0.98 | 1.62 | 0.075 |
| Model 3: *Model 2 + demographics & genetics* [b] | 1.19 | 0.16 | 0.92 | 1.54 | 0.192 |
| Model 4: *Fully Adjusted* [c] | 0.86 | 0.12 | 0.66 | 1.13 | 0.280 |
| **Adjustments** | **Long Sleep** | | | | |
| | **RRR** | **SE** | **95% CI** | | ***p*** |
| ***Moderate-risk* Profile** | | | | | |
| Model 1: *Unadjusted* | 1.55 | 0.48 | 0.85 | 2.84 | 0.152 |
| Model 2: *Model 1 + baseline biomarkers* [a] | 1.43 | 0.45 | 0.77 | 2.66 | 0.254 |
| Model 3: *Model 2 + demographics & genetics* [b] | 1.16 | 0.37 | 0.62 | 2.16 | 0.650 |
| Model 4: *Fully Adjusted* [c] | 1.05 | 0.34 | 0.55 | 1.98 | 0.893 |
| ***High-risk* Profile** | | | | | |
| Model 1: *Unadjusted* | 3.52 | 1.03 | 1.98 | 6.24 | <0.001 |
| Model 2: *Model 1 + baseline biomarkers* [a] | 2.78 | 0.94 | 1.43 | 5.41 | 0.003 |
| Model 3: *Model 2 + demographics & genetics* [b] | 2.02 | 0.70 | 1.02 | 3.98 | 0.043 |
| Model 4: *Fully Adjusted* [c] | 1.48 | 0.53 | 0.73 | 2.99 | 0.277 |

Notes: The *low-risk* group is the reference; RRR = relative risk ratio; SE = standard errors; CI = confidence interval; *p* = significance value.
a Baseline immune and neuroendocrine profiles.
b Demographic and genetic variables: age; sex; 10 principal components (PCs); C-reactive Protein (CRP) polygenic score (PGS); White Blood Cell Counts (WBCC) PGS; Insulin Growth Factor-1 (IGF-1) PGS; [Hair] Cortisol PGS; Sleep Duration PGS.
c All variables: Baseline immune and neuroendocrine profiles; age; sex; 10 PCs; CRP PGS; WBCC PGS; IGF-1 PGS; Cortisol PGS; education; wealth; occupational social status; smoking status; alcohol consumption; physical activity; mobility; limiting longstanding illness; health (i.e., chronic lung disease; coronary heart disease; abnormal heart rhythm; heart murmur; congestive heart failure; angina; hypertension; diabetes; cancer; Parkinson's; Alzheimer's; dementia; asthma; arthritis; osteoporosis; psychiatric disorder); BMI.

***Table S10.*** **Longitudinal associations between suboptimal sleep durations at less stringent thresholds (≤6 hr; >6-<8 hr; and ≥8 hr) and immune and neuroendocrine profile membership**

| Adjustments | Short Sleep | | | | |
|---|---|---|---|---|---|
| | **RRR** | **SE** | **95% CI** | | ***p*** |
| ***Moderate-risk* Profile** | | | | | |
| Model 1: *Unadjusted* | 1.21 | 0.09 | 1.05 | 1.39 | 0.010 |
| Model 2: *Model 1 + baseline biomarkers* [a] | 1.13 | 0.09 | 0.98 | 1.31 | 0.098 |
| Model 3: *Model 2 + demographics & genetics* [b] | 1.08 | 0.08 | 0.93 | 1.25 | 0.329 |
| Model 4: *Fully Adjusted* [c] | 0.99 | 0.08 | 0.85 | 1.15 | 0.885 |
| ***High-risk* Profile** | | | | | |
| Model 1: *Unadjusted* | 1.46 | 0.12 | 1.24 | 1.71 | <0.001 |
| Model 2: *Model 1 + baseline biomarkers* [a] | 1.26 | 0.12 | 1.05 | 1.52 | 0.014 |
| Model 3: *Model 2 + demographics & genetics* [b] | 1.20 | 0.12 | 0.99 | 1.46 | 0.058 |
| Model 4: *Fully Adjusted* [c] | 0.97 | 0.10 | 0.79 | 1.18 | 0.736 |
| **Adjustments** | **Long Sleep** | | | | |
| | **RRR** | **SE** | **95% CI** | | ***p*** |
| ***Moderate-risk* Profile** | | | | | |
| Model 1: *Unadjusted* | 1.10 | 0.16 | 0.83 | 1.45 | 0.507 |
| Model 2: *Model 1 + baseline biomarkers* [a] | 1.00 | 0.15 | 0.75 | 1.34 | 0.982 |
| Model 3: *Model 2 + demographics & genetics* [b] | 0.88 | 0.13 | 0.65 | 1.17 | 0.367 |
| Model 4: *Fully Adjusted* [c] | 0.85 | 0.13 | 0.63 | 1.14 | 0.282 |
| ***High-risk* Profile** | | | | | |
| Model 1: *Unadjusted* | 2.09 | 0.30 | 1.58 | 2.76 | <0.001 |
| Model 2: *Model 1 + baseline biomarkers* [a] | 1.61 | 0.27 | 1.16 | 2.24 | 0.005 |
| Model 3: *Model 2 + demographics & genetics* [b] | 1.31 | 0.23 | 0.94 | 1.84 | 0.116 |
| Model 4: *Fully Adjusted* [c] | 1.18 | 0.21 | 0.83 | 1.67 | 0.354 |

Notes: The *low-risk* group is the reference; RRR = relative risk ratio; SE = standard errors; CI = confidence interval; *p* = significance value.

a Baseline immune and neuroendocrine profiles.

b Demographic and genetic variables: age; sex; 10 principal components (PCs); C-reactive Protein (CRP) polygenic score (PGS); White Blood Cell Counts (WBCC) PGS; Insulin Growth Factor-1 (IGF-1) PGS; [Hair] Cortisol PGS; Sleep Duration PGS.

c All variables: Baseline immune and neuroendocrine profiles; age; sex; 10 PCs; CRP PGS; WBCC PGS; IGF-1 PGS; Cortisol PGS; education; wealth; occupational social status; smoking status; alcohol consumption; physical activity; mobility; limiting longstanding illness; health (i.e., chronic lung disease; coronary heart disease; abnormal heart rhythm; heart murmur; congestive heart failure; angina; hypertension; diabetes; cancer; Parkinson's; Alzheimer's; dementia; asthma; arthritis; osteoporosis; psychiatric disorder).

***Table S11.*** **The longitudinal role of PGS for suboptimal sleep durations in immune and neuroendocrine profile membership**

| **Adjustments** | **PGS for Short Sleep** | | | | |
|---|---|---|---|---|---|
| | **RRR** | **SE** | **95% CI** | | ***p*** |
| ***Moderate-risk* Profile** | | | | | |
| Model 1: *Unadjusted* | 1.01 | 0.03 | 0.95 | 1.08 | 0.818 |
| Model 2b: *Model 1 + demographics & genetics* [a] | 0.99 | 0.04 | 0.92 | 1.07 | 0.831 |
| ***High-risk* Profile** | | | | | |
| Model 1: *Unadjusted* | 1.04 | 0.04 | 0.97 | 1.12 | 0.300 |
| Model 2b: *Model 1 + demographics & genetics* [a] | 1.00 | 0.05 | 0.91 | 1.10 | 0.992 |
| **Adjustments** | **PGS for Long Sleep** | | | | |
| | **RRR** | **SE** | **95% CI** | | ***p*** |
| ***Moderate-risk* Profile** | | | | | |
| Model 1: *Unadjusted* | 0.99 | 0.03 | 0.93 | 1.05 | 0.694 |
| Model 2b: *Model 1 + demographics & genetics* [a] | 0.97 | 0.03 | 0.91 | 1.04 | 0.439 |
| ***High-risk* Profile** | | | | | |
| Model 1: *Unadjusted* | 1.06 | 0.04 | 0.99 | 1.15 | 0.097 |
| Model 2b: *Model 1 + demographics & genetics* [a] | 1.03 | 0.05 | 0.94 | 1.13 | 0.555 |

Notes: The *low-risk* group is the reference; RRR = relative risk ratio; SE = standard errors; CI = confidence interval; *p* = significance value.
a Demographic and genetic variables: age; age$^2$; sex; 10 principal components (PCs).

**Supplementary Materials 1**

**C-reactive Protein.** High-sensitivity plasma CRP (mg/L) was assayed using the N Latex CRP mono Immunoassay on the Behring Nephelometer II analyser (Dade Behring, Milton Keynes, UK). Intra and inter-assay coefficients of variation were <2%. The lower detection limit of the assay was 0.2 mg/L. CRP values >20 mg/L were excluded from analyses (*n*=116), as these were taken to reflect acute inflammatory processes rather than chronic inflammation (Hamilton et al., 2021). CRP was treated as continuous, with higher values indicating greater levels of inflammation.

**Fibrinogen.** Plasma fibrinogen (g/L) was analysed using a modification of the Clauss thrombin clotting method on the Organon Teknika MDA 180 coagulation analyser (Organon Teknika, Durham, USA). Intra and inter-assay coefficients of variation were <7%. The lower detection limit of the assay was 0.5 g/L. Fibrinogen was treated as continuous, with higher values indicating greater levels of inflammation.

**Leukocytes (White Blood Cell Counts [WBCC]).** WBCC was analysed as continuous counts per 109/L; measured on a haematology-automated analyser (Abbott Diagnostics Cell-Dyn 4000 and Sysmex XE), with higher values indicating greater levels of inflammation.

**Insulin-like Growth Factor-1.** Serum IGF-1 (nmol/L) was measured using the DPC Immulite 2000 method, by an electrochemiluminescent immunoassay on IDS ISYS Analyser. Inter and intra-assay coefficients of variation were <14%. IGF-1 was treated as continuous, with lower values indicating greater neuroendocrine activity.

**Hair Cortisol.** Hair strands ~3cm, weighing ~10mg were collected from the posterior vertex, as close to the scalp as possible. Assuming an average hair growth of ∼1cm per month,[16] the hair segment closest to the scalp is thought to provide a measure of the average cortisol output over the preceding three months prior to sampling. Exclusion criteria for hair sampling included pregnancy, breastfeeding, select scalp conditions, having <2cm of hair length, and an inability keep head still. Hair cortisol concentrations were analysed at the Technische Universität Dresden (Germany). Cortisol levels were assayed using high performance liquid chromatography-mass spectrometry (LC/MS) following a standard wash and steroid extraction procedure,[17] and were expressed in pg/mg. Data was log-transformed, as the distribution was positively skewed.

**Supplementary Materials 2**

**Directed Acyclic Graph (DAG).** Covariates were selected *a priory* through a DAG (Figure S2). The DAG serves as validation for the proper parameterisation of the models, to reduce overadjustment bias, and to ensure the adherence of assumptions, *inter alia* homoscedasticity and an absence of interactions (Nilsson et al., 2021; Van Zwieten et al., 2022). It is important to note that while the DAG identifies the presence of bias, it does not explicitly specify the type nor the magnitude of the bias, whether there are competing biases, or whether the observed bias is clinically meaningful (Lipsky & Greenland, 2022).

**Supplementary Materials 3**

**PGS Derivation for Sleep Duration, and Immune and Neuroendocrine Biomarkers**

The genome-wide genotyping, funded by the Economic and Social Research Council (ESRC), was performed at University College London (UCL) Genomics in 2013-2014 using the llumina HumanOmni2.5 BeadChips (HumanOmni2.5-4v1, HumanOmni2.5-8v1.3), which measures ~2.5 million markers that capture the genomic variation down to 2.5% minor allele frequency (MAF).

**Genetics Data Quality Control.** The methods employed for quality control of genomic data in the ELSA study are those outlined by the Health and Retirement Study (HRS).[1] This was done to harmonise the research across the age-related longitudinal studies by adopting a consistent methodology. Single-nucleotide polymorphism (SNPs) were excluded if they were non-autosomal, MAF was <1%, if more than 2% of genotype data were missing and if the Hardy-Weinberg Equilibrium $p<10^{-4}$. Samples were removed based on call rate (<0.99), heterozygosity, relatedness and if the recorded sex phenotype was inconsistent with genetic sex. To identify ancestrally homogenous analytic samples the ELSA genomic samples use a combination of both self-reported ethnicity and analyses of genetic ancestry. To improve genome coverage, we imputed untyped quality-controlled genotypes to the Haplotype Reference Consortium[2] using the University of Michigan Imputation Server.[3] Post-imputation, we kept variants that were genotyped or imputed at INFO>0.80, in low linkage disequilibrium ($R^2<0.1$) and with Hardy-Weinberg Equilibrium $p$-value$>10^{-5}$. After the sample quality control 7179780 variants were retained for further analyses. To account for potentially biasing ancestry differences in genetic structures, a principal components (PCs) analysis was conducted, retaining the top 10 PCs,[4] which were subsequently used to adjust for possible population stratification in the association analyses.[4,5] Genetic ancestry was estimated via comparison of participants' genotypes to global reference populations using principal component analyses (PCA). Because PCA allows examining population structure in a cohort by determining the average genome-wide genetic similarities of individual samples, derived principal components (PCs) can be used to group individuals with shared genetic ancestry, to identify outliers, and as covariates, to reduce false positives due to population stratification. Although up to 98% of the ELSA participants self-described to be of European cultural background, PC highlighted the present of ancestral admixture in $n$=65 (0.9%) individuals (implying these individuals had ancestors from two or more populations. Even though this type of labelling of ancestral populations oversimplifies the complexity of human genetic variation, accounting for systematic differences in allele frequencies is necessary for genetic analyses. Therefore, these participants with ancestral admixture were removed from the analyses. The final sample includes all self-reported European participants that had PC loadings within ± (a standard deviation) from the mean for eigenvectors one. PCs were then re-calculated to further account for population stratification. Therefore, our analytic sample included the full ELSA sample that provided genetic samples and passed quality control. We further utilized the PCs for adjusting for possible population stratification in the association analyses.

**Polygenic risk scores (PGS).** PGSs for sleep duration in ELSA[6] using the GWAS summary statistics perfumed using the data from the UK Biobank.[7] Sleep duration was a self-reported phenotype where participants were asked the number of hours sleep (in one hour increments) that they typically get in a 24hr period, including naps. Participant DNA was genotyped on two arrays, UK BiLEVE and UKB Axiom, with >95% common content. Genotypes for 152,736 samples passed sample quality control (~99.9% of total samples). Before imputation, 806,466 SNPs passed quality control in at least one batch (>99% of the array content). Imputation of autosomal SNPs was performed to a merged reference panel comprising the Phase 3 1000 Genomes Project and UK10K panels. Genetic association analysis for autosomes was performed in SNPTEST with the 'expected' method using an additive genetic model adjusted for age, sex, 10 principal components and genotyping array. The PGS distribution was normal, with a mean of 5,610.20 ($_{SE}$0.18;

$_{range}$5552.6-5665.2). A total of 948,331 SNPs overlapped with the ELSA genetic database with the GWAS summary statistics and were included in the PGS for this phenotype.

PGS for C-reactive protein in ELSA[6] was contracted using the results from UK Biobank (UKB) genome-wide association studies (GWAS) [8], based on 427,367 individuals. The GWAS identified 49,164 genetic loci at a genome-wide significance of $p<5\times10^{-8}$. Linear Mixed Model (LMM) regression using BOLT-LMM version 2.343 was performed on CRP levels in UKB. This model accounts for cryptic relatedness within the sample. An additive genetic model was used for all 8.9 million measured and imputed genetic variants. The model was adjusted for age, sex, UKB array (UKB vs UK BiLEVE to account for the different genotyping chips) and 40 genetic principal components. Serum CRP levels (mg/l) was measured by immunoturbidimetry. CRP levels were transformed using natural log and the resulting range was from -2.53-4.38, excluding individuals with extreme values ±4 from the mean. 1.8% of the sample was removed because they had an autoimmune disorder and were on immunosuppressive drugs.

PGS for WBCC in ELSA[6] was calculated using summary statistics from GWAS meta-analyses that included data from the UKB and a largescale international collaborative effort, including data for 563,085 European ancestry participants. There were 27,090,932 genetic loci at a genome-wide significance of $p<5\times10^{-8}$, with 5,106 new genetic variants independently associated with 29 blood cell phenotypes covering a range of variation impacting hematopoiesis. The WBCC phenotype (109/L), an aggregate number of white blood cells per unit volume of blood, is one of several quantitative clinical laboratory measures that together reflect hematopoietic progenitor cell production, hemoglobin synthesis, maturation, release from the bone marrow, and clearance of mature or senescent blood cells from the circulation [9]. Raw phenotypes were regressed on age, $age^2$, sex, principal components, and cohort specific covariates. WBCC related traits were $log^{10}$ transformed before regression modeling. Residuals from the modeling were obtained and then inverse normalised. The cohort level association analyses were conducted using a linear mixed effects model to account for known or cryptic relatedness (e.g., BOLT-LMM, EPACTS https://github.com/statgen/EPACTS and rvtests with the additive genetic model). Linear mixed effects models have been shown to effectively account for both population structure and inter-individual relatedness within the UK Biobank cohort, along with having increased discovery power over simple linear regression with principal components.

PGS for IGF-1 was calculated using summary statistics from GWAS that included 10,280 men and women in the analyses, comprising 1712 participants in the Cardiovascular Health Study (CHS), 3507 in the Framingham Heart Study (FHS), 1607 participants in the Cooperative Research in the Region of Augsburg (KORA) study and 3454 in the Study of Health in Pomerania (SHIP) [10]. Analyses of SNP associated with IGF-1 concentrations revealed that rs700752 was associated with IGF-I concentrations ($p$=4.9×10$^{-9}$), but this was attenuated (meta-analysis $p$=0.038) after adjustment for IGFBP-3 concentrations. Three additional SNPs achieved $p$<10$^{-6}$ in relation to IGF-I concentrations: rs2153960 on chromosome 6q21, MAF=0.31, $p$=5.1×10$^{-7}$; rs1245541 on chromosome 10q22.1, MAF=0.39, $p$=5.0×10$^{-7}$; rs7780564 on chromosome 7p21.3, MAF=0.45, $p$=3.9×10$^{-7}$.

PGS for Morning Plasma Cortisol in ELSA[6] was contracted using the results from the CORtisol NETwork (CORNET) consortium, which undertook the GWAS meta-analysis for plasma cortisol in 12,597 White participants from 11 western European population-based cohorts and replicated their results in 2,795 participants from three independent cohorts.[11] Cortisol was measured by immunoassay in blood samples collected from study participants between 07:00h and 11:00h. Each study performed single marker association tests, and study-specific linear regression models which used z-scores of log-transformed cortisol, additive SNP effects, and were adjusted for age and sex (model 1); age, sex, and smoking (model 2); or age, sex, smoking and body mass index (model 3). Imputation of the gene-chip results used the HapMap CEU population, build 36. The results indicate that <1% of variance in plasma cortisol is accounted for by genetic variation in a single region of chromosome 14. The CORNET GWAS summary statistics for this phenotype contained 2,660,191 SNPs; of these, 837,709 SNPs overlapped with the ELSA genetic database and were included in the PGS for Morning Plasma Cortisol phenotype.

**Supplementary Materials 4**

**Multiple Imputation.** Although missingness reached 61.0% (Table S1), multiple imputation can reduce bias even when the proportion of missingness is substantial (Madley-Dowd et al., 2019). Thus, for a better powered sample, for greater precision, and to preclude the possibility of bias in the complete case analyses (Sterne et al., 2009), we imputed missing data on exposures, covariates, and outcomes, with the exclusion of wholly absent biological and genetic data. Imputation was performed using missForest (R v.4.2.0: RStudio v.2022.02.2); a Random Forests algorithm-based machine learning imputation method. Random-forests is non-parametric, it works with high-dimensional data, and it has a built-in feature selection that evaluates entropy and information gain, so it is robust to noisy data and multicollinearity. In the presence of nonlinearity and interactions missForest outperformed prominent imputation methods, such as multivariate imputation by chained equations and k-nearest neighbours in all metrics (Stekhoven & Bühlmann, 2012). Given that socioeconomic and health-related variables are the main drivers of attrition in ELSA (Steptoe et al., 2013), the assumption that missingness was at random (MAR) was likely to be met in our analysis since these variables were included in the imputation models. Imputed and observed data were homogenous (Table S1) and the earlier use of the study data have shown that results deriving from imputed data aligned with those from complete case analyses (Hamilton, Iob, et al., 2023; Hamilton, Steptoe, et al., 2023).

**Supplementary Materials 5**

**Latent Profile Analysis (LPA).** Outcomes of immune and neuroendocrine biomarkers were entered into the LPA, which included high-sensitivity plasma C-reactive protein (CRP; mg/L), plasma fibrinogen (Fb; g/L), serum insulin-like growth factor-1 (IGF-1; mmol/L) and hair cortisol (cortisol; pg/mg). A stepwise approach was taken to identify the optimal number of latent profiles; starting with a single-profile model, additional profiles were added to improve model fit. The number of profiles was determined on the basis of the Akaike information criterion (AIC) (Akaike, 1998), Bayesian information criterion (BIC) (Schwarz, 1978), and adjusted Bayesian information criterion (aBIC) (Bozdogan, 1987). The information criteria and the likelihood ratio tests indicated the goodness of fit of different latent profile models, with the best model being the one with the lowest AIC, BIC, and aBIC values. The entropy statistic that provides the quality of the classification model, and the average posterior probabilities for each latent profile, indicating profile membership classification errors, were also taken into account (Celeux & Soromenho, 1996). The closer to 1 these indicators were, the better the classification quality (Morin et al., 2016). A common cut-off point for posterior probabilities is 0.70 or above (Nagin, 2009). An entropy of 0.80 or greater indicates clear profile separation (Kamata et al., 2018). Every profile must contain ≥5% of participants and the profiles must be of good theoretical interpretability (Herle et al., 2020). Once the number of latent profiles was established, each individual within the sample was assigned to a cluster for which they had the largest posterior probability, indicating the most likely affiliation.
